# Anatomy of the first six months of COVID-19 Vaccination Campaign in Italy

**DOI:** 10.1101/2021.11.24.21266820

**Authors:** Nicolò Gozzi, Matteo Chinazzi, Jessica T. Davis, Kunpeng Mu, Ana Pastore y Piontti, Marco Ajelli, Nicola Perra, Alessandro Vespignani

## Abstract

We analyze the effectiveness of the first six months of vaccination campaign against SARS-CoV-2 in Italy by using a computational epidemic model which takes into account demographic, mobility, vaccines, as well as estimates of the introduction and spreading of the more transmissible Alpha variant. We consider six sub-national regions and study the effect of vaccines in terms of number of averted deaths, infections, and reduction in the Infection Fatality Rate (IFR) with respect to counterfactual scenarios with the actual non-pharmaceuticals interventions but no vaccine administration. Furthermore, we compare the effectiveness in counterfactual scenarios with different vaccines allocation strategies and vaccination rates. Our results show that, as of 2021/07/05, vaccines averted 29, 350 (*IQR*: [16, 454 − 42, 826]) deaths and 4, 256, 332 (*IQR*: [1, 675, 564 − 6, 980, 070]) infections and a new pandemic wave in the country. During the same period, they achieved a −22.2% (*IQR*: [−31.4%; −13.9%]) reduction in the IFR. We show that a campaign that would have strictly prioritized age groups at higher risk of dying from COVID-19, besides frontline workers, would have implied additional benefits both in terms of avoided fatalities and reduction in the IFR. Strategies targeting the most active age groups would have prevented a higher number of infections but would have been associated with more deaths. Finally, we study the effects of different vaccination intake scenarios by rescaling the number of available doses in the time period under study to those administered in other countries of reference. The modeling framework can be applied to other countries to provide a mechanistic characterization of vaccination campaigns worldwide.

## Introduction

After almost a year marked by the implementation of non-pharmaceutical interventions (NPIs) [1–5] and enormous losses in terms of human lives and socioeconomic disruptions, on the 27^*th*^ of December, 2020, simultaneously with other European countries, the first dose of vaccine against COVID-19 was administered in Italy [6]. The vaccine rollout proceeded prioritizing healthcare personnel, care facilities residents, and 80+ individuals. Unfortunately, as in many other countries, the success of the COVID-19 vaccination campaign was hindered by several obstacles. Delays in deliveries from suppliers, temporary suspensions and changes in the administration protocol of the AstraZeneca vaccine, and the logistical issues linked to the delivery of millions of doses over the national territory are just some examples. Furthermore, the vaccine rollout and its effect on the pandemic were challenged by the March 2021 epidemic surge due to the spread of the more transmissible SARS-CoV-2 Alpha variant (Pango lineage B.1.1.7), first detected in the United Kingdom in September 2020 [7–12]. In Italy, the first Alpha case was identified in late December 2020. By week 4 of 2021 the variant was responsible for more than 50% of newly reported cases, and for more than 80% by week 12 [13].

In this complex epidemiological landscape, it is extremely important to provide a characterization of the effects of the vaccination program and to which extent they contributed to a decrease in the number of newly reported cases and deaths. In the United Kingdom it has been estimated that the vaccines averted around 30^*′*^000 additional deaths during the first six months of 2021 [14] and saving around 279^*′*^000 lives in the United States during the same period [15]. In the context of Italy, optimal allocation of vaccines [16, 17] and the potential combined effect of NPIs and rollout on epidemic scenarios [18] have been explored. A national level study [19], explored the impact of the vaccination program in Italy and evaluate possible prospects for reopening the society.

Here, we develop a computational multi-strain epidemic model able to provide a detailed sub-national characterization of the first six months of the Italian COVID-19 vaccination campaign. We inform the model with real data on mobility changes [20] and policy interventions [21] capturing the variations in contacts modulated by NPIs. We use a mechanistic approach based on international travel flows to characterize the Alpha variant introductions [22, 23]. Data on vaccines rollout come from official sources of the Italian Government [24] and consider separately the four vaccines currently authorized in Italy: Pfizer/Biontech, Moderna, Vaxzevria (Astrazeneca), and Janssen (Johnson & Johnson). This data provides the number of daily inoculations by age, priority group, geographical area, and vaccine supplier. The model is calibrated on reported weekly deaths over the period 2020/09/01 - 2021/07/05 separately for different NUTS1 regions through an Approximate Bayesian Computation method [25].

We estimate that vaccines averted 29, 350 (*IQR*: [16, 454 − 42, 826]) deaths and 4, 256, 332 (*IQR*: [1, 675, 564−6, 980, 070]) infections between 2020/12/27 and 2021/07/05 with respect to a counterfactual scenario without vaccines and the actually implemented NPIs. In the same period, they contributed to an overall reduction of the Infection Fatality Rate (IFR) by − 22.2% (*IQR*: [− 31.4%; − 13.9%]). We also estimate that even even with stringent additional NPIs, the absence of vaccines would have led to 12*K*+ more deaths and 540*K*+ infections. Furthermore, we study counterfactual scenarios in which we either assign vaccine in a very strict decreasing age order or first to 20-49 age groups. We estimate that 31, 786 (*IQR*: [19, 115 − 44, 733]) deaths would have been averted in the first scenario and only 21, 440 (*IQR*: [8, 006−35, 429]) when prioritizing younger age groups. Nonetheless, the second strategy would have implied less infections. We also explore the effects of different rollout speeds applying the vaccination rates observed in the United Kingdom (UK) and the United States (US). As expected, when more doses are available (vaccination rates of UK or US), additional benefits are observed in terms of averted deaths and infections. Interestingly, we find that not only the number of vaccines but also the timing of availability is an important factor determining the outcome of the vaccination campaign. For instance, a vaccination rate timeline similar to the US one would have resulted in an additional ∼ 45% averted deaths (median: 42, 578, *IQR*: [29, 768 − 55, 835]).

The presented modeling framework is a general tool for the mechanistic study of counterfactual scenarios evaluating, and informing the design of vaccination campaigns, and can be extended to additional countries depending on data availability.

## Results

We adopt a SLIR-like compartmentalization setup with the addition of specific compartments to account for vaccination and the emergence of a more transmissible virus strain. The model includes the age-stratification of the population and of their contacts. Specifically, the population is divided into ten age groups and the contacts between them are defined by a country-specific contacts matrix *C* from Ref. [26]. Variations in contacts induced by non-pharmaceutical interventions at workplaces and in the community settings are modelled considering data from the COVID-19 Community Mobility Report published by Google [20]. We account for restrictions in schools using the Oxford COVID-19 Government Response Tracker [21] and the timeline of government interventions. The model includes a seasonal modulation to account for variations in factors such as humidity and temperature that can influence transmissibility [23, 27]. As a way to include a second, more transmissible virus strain, the compartmental structure is extended with specific Latent and Infectious compartments. We refer the reader to the Material and Methods section and the Supplementary Information for further details about the epidemic model as well as for a sensitivity analysis around the choice of parameters presented below.

To capture geographical heterogeneities in vaccines’ administration, spreading of the virus, and variant’s importations our model is run for each NUTS1 region in Italy. In addition, we split the NUTS1 region Isles into its two NUTS2 territories, Sicily and Sardinia. As a result, we model six different areas: North West, North East, Center, South, Sicily and Sardinia. The number of individuals in different age groups for each region is taken from the official census [28]. We use official sources for the epidemiological data [29]. As detailed in the Material and Methods section, the model is calibrated separately for each of the six regions using an Approximate Bayesian Computation (ABC) technique [25, 30]. We set the calibration period to 2020/09/01-2021/07/05 and we use weekly deaths as output quantity. The free parameters are the transmissibility *β*, the delay in deaths reporting Δ, the modulation of seasonality *α*_*min*_, and the initial conditions for the number of individuals in each compartment.

We model the vaccines rollout in Italy from official data [24] which provides the number of daily doses (1^*st*^ and 2^*nd*^) divided by age groups, risk category, supplier, and region. In Fig. 1A we plot the number of 1^*st*^ and 2^*nd*^ doses administered daily in the country as reported by official sources [24]. The first batch of Pfizer/BioNTech vaccines received in late December 2020 was administered mainly to healthcare workers and care facilities residents. These individuals received the 2^*nd*^ dose in the last two weeks of January. This explains the very low number of 1^*st*^ doses given in this time range. Similarly, the decline observed since early June 2021 is due to the high number of 2^*nd*^ doses administered during this period. As of 2021/07/05, 58.1% of the total population received at least one dose, while 32.7% received two, with low variability across different regions (see Fig. 1B). In Fig. 1C we show the average age of the vaccinated in time. We see the following trend: in January/February 2021 the average age is around 50, this is due to the vaccinations of healthcare personnel; after, the campaign proceeded prioritizing mainly the elderly as shown by the increase of average age; finally, from mid-April average age of the vaccinated declines since the rollout was extended to younger age brackets. The grey lines indicate the average age of the vaccinated in the different regions considered. As we can see, they do not diverge significantly from the national average. Finally, in Fig. 1D we show the total number of (first) doses administered by vaccine suppliers. We see that most of the people received the Pfizer/BioNTech vaccines (68.9%), followed by Vaxzevria (AstraZeneca) (18.4%), Moderna (9.0%), and Janssen (Johnson&Johnson) (3.7%).

**Figure 1:**
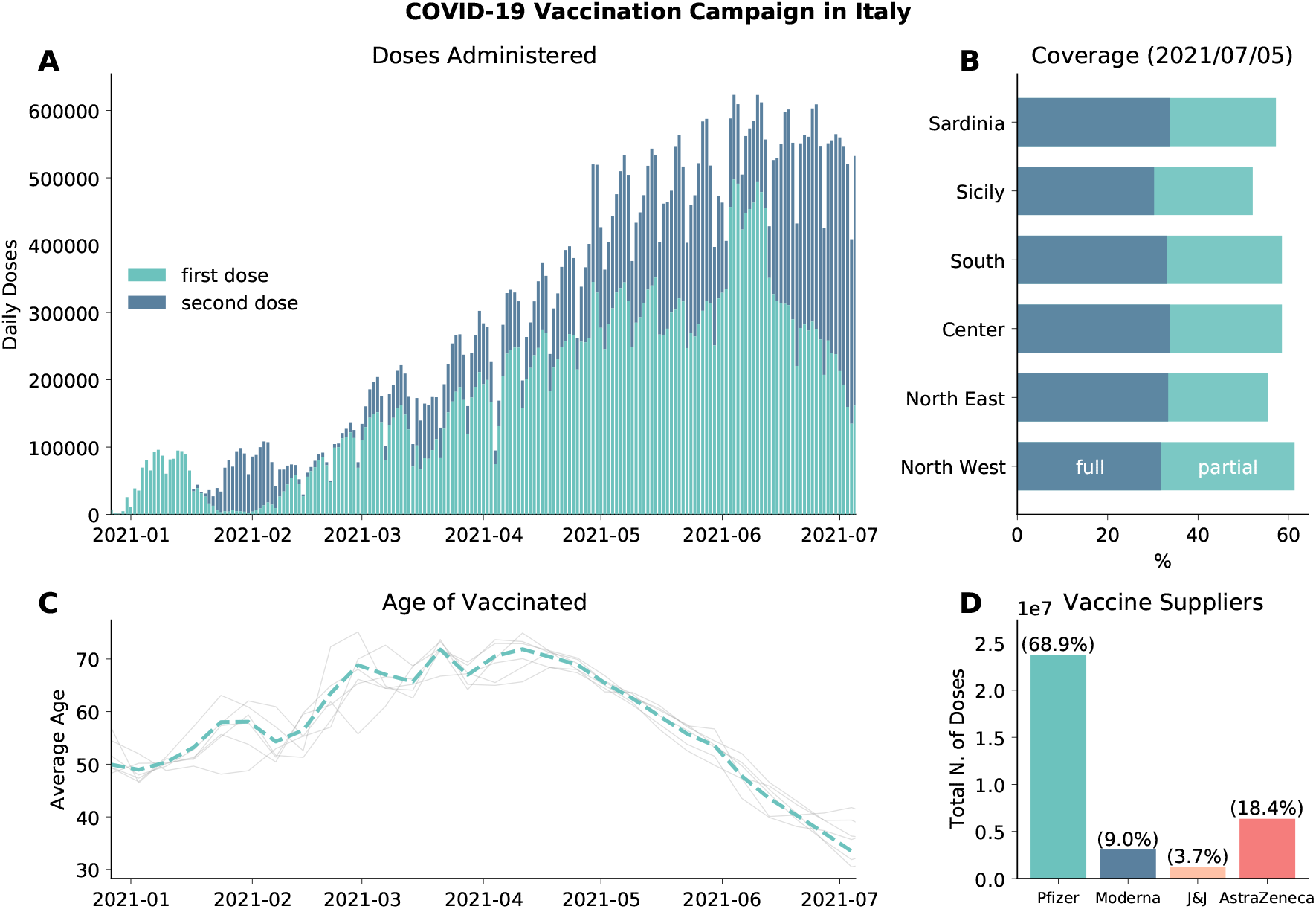
Vaccination campaign in Italy. A) Number of daily first and second administered daily in the country. B) Percentage of total population vaccinated with one/two dose in different regions considered. C) Average age of those who received the first dose of vaccines (dashed line). Grey lines indicate the average age of vaccinated in different regions considered. D) Total number of first doses administered between 2020/12/27 and 2021/07/05 by vaccine supplier.

Our model explicitly accounts for the emergence and spread of the more transmissible SARS-CoV-2 variant Alpha. The introductions of this lineage in the different regions are estimated with the Global Epidemic and Mobility model (GLEAM [22, 23, 31]) using actual origin-destination data during the period 2020/09/01-2020/12/31 (see details in the Material and Methods section). In Fig. 2A we show the estimated date of dominance yield by the model calibration, defined as the first day in which Alpha variant was responsible for at least 50% of the infections. Across the different regions, we estimate that Alpha became dominant within the first two weeks of March, 2021.

**Figure 2:**
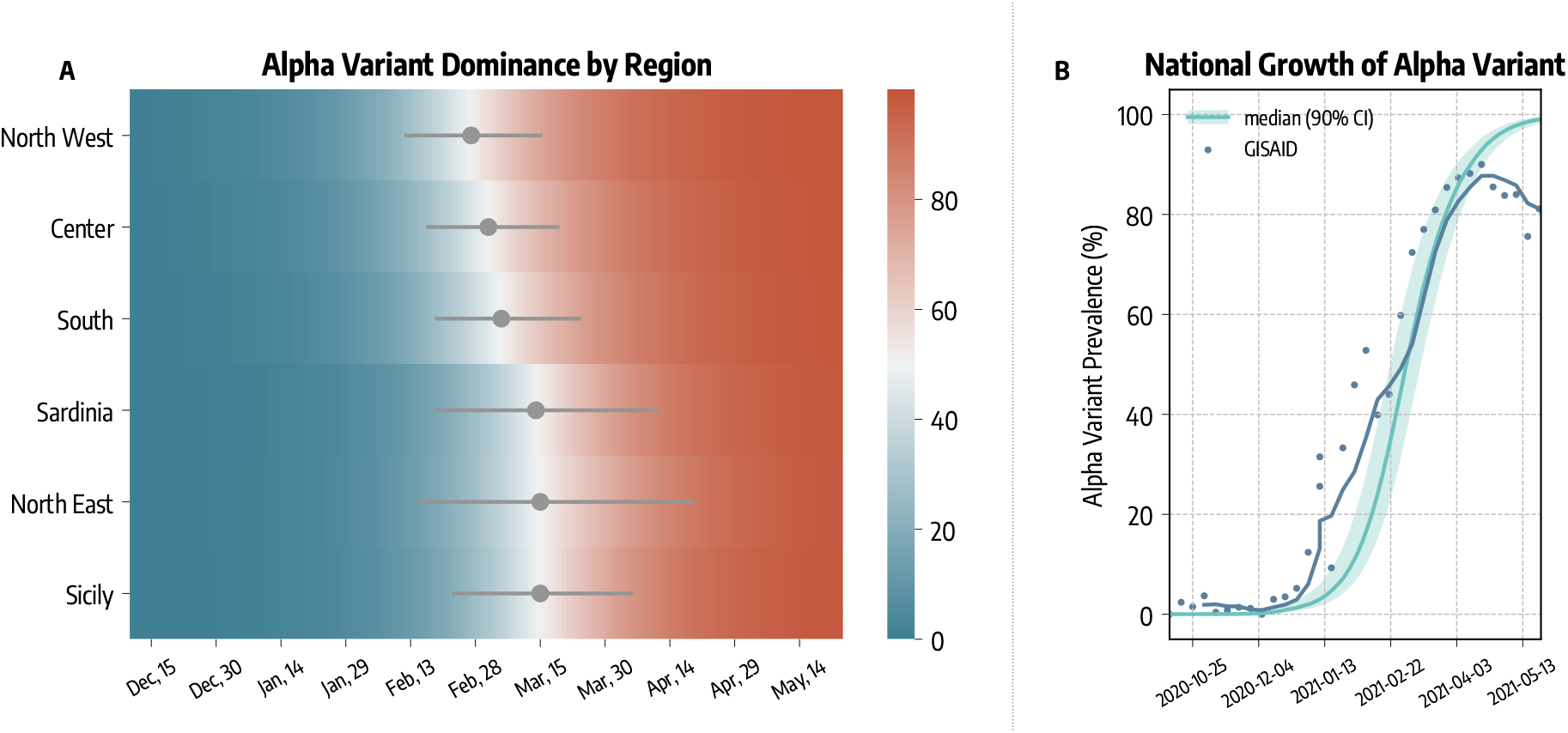
Alpha Variant Spreading and Dominance. A) Estimated date of dominance of the Alpha variant in different regions. B) Comparison between estimated and reported national prevalence of Alpha variant.

In Fig. 2B we compare the estimated national Alpha variant prevalence (median and 90% CI) with available genomic data from GISAID [13, 32]. Our modeling approach is able to reproduce the national growth of Alpha variant. The correlation between the simulated and real prevalence (Pearson *ρ* = 0.84, *p <* 0.001) is remarkable (weighted mean absolute percentage error *wMAPE* = 0.34). We acknowledge a deviation between simulated and real prevalence by late April. This is due to the emergence of the Delta variant, which has been estimated to be more transmissible than Alpha [33] and that we do not include in our model.

### Averted Deaths and Infections

In Fig. 3A we show the estimated number of COVID-19 deaths and infections averted by vaccines. To obtain these quantities, we first calibrate the model using real data on mobility, policy interventions, epidemic evolution, and vaccines rollout. Then, we compare the number of projected deaths and infections in simulations with vaccines administered and without.

**Figure 3:**
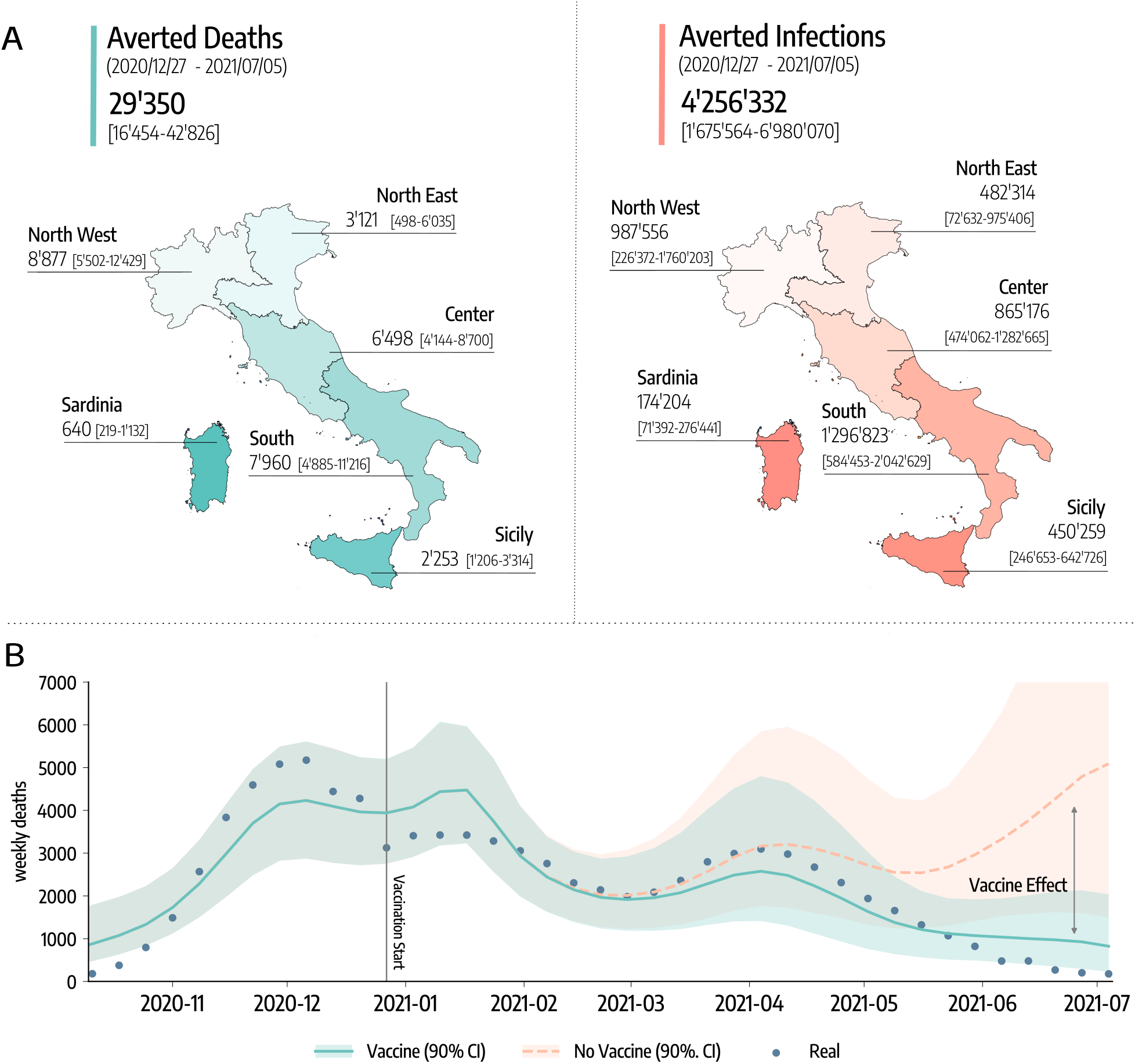
Averted deaths and infections. A) Estimated averted deaths and infections in different regions considered with respect to a baseline without vaccines. Median and interquartile (*IQR*) range are reported. B) Number of weekly deaths at the country level as reported by official surveillance and as estimated by our model with and without vaccines rollout (median and 90% confidence intervals displayed).

The results suggest that vaccines avoided 29, 350 (*IQR*: [16, 454−42, 826]) deaths between 2020/12/27-2021/07/05. This is about 50% of the number of deaths reported in the country during the same period. If we look at the different regions considered, 8, 877 (*IQR*: [5, 502 − 12, 429]) deaths were averted in North West, 3, 121 (*IQR*: [498 − 6, 035]) in North East, 6, 498 (*IQR*: [4, 144 − 8, 700]) in Center, 7, 960 (*IQR*: [4, 885−11, 216]) in South, 2, 253 (*IQR*: [1, 206−3, 314]) in Sicily, and 640 (*IQR*: [219−1, 132]) in Sardinia. To give a better idea, these numbers (medians) corresponds to the following percentages of the total number of deaths observed in the regions during the same period: 58% in North-West, 20% in North East, 60% in Center, 65% in South, 63% in Sicily, and 77% in Sardinia.

Similarly, 4, 256, 332 (*IQR*: [1, 675, 564−6, 980, 070]) infections were avoided in the country, divided into the different regions as follows: 987, 556 (*IQR*: [226, 372 − 1, 760, 203]) in North West, 482, 314 (*IQR*: [72, 632 − 975, 406]) in North East, 865, 176 (*IQR*: [474, 062 − 1, 282, 665]) in Center, 1, 296, 823 (*IQR*: [584, 453 − 2, 042, 629]) in South, 450, 259 (*IQR*: [246, 653 − 642, 726]) in Sicily, and 174, 204 (*IQR*: [71, 392 − 276, 441]) in Sardinia.

Furthermore, our results suggest that vaccines prevented an additional COVID-19 wave. In Fig. 3B we show the estimated number of weekly deaths (median and 90% CI) at the national level in the simulations with and without vaccines. The two curves start to visibly diverge in mid-March, around the peak of the wave of infections led by the emergence and dominance of the Alpha variant. Interestingly, the difference between the two curves becomes even bigger by late April, when some restrictions were partially eased in the country. We estimate that, in absence of the vaccines, this reopening would led to a rapid resurgence in infections and deaths, reaching a peak higher than those observed in January and March 2021. We note how this result should be interpreted carefully. Indeed, an hypothetical upturn in fatalities would have likely led to countermeasures, a u-turn in the reopening timeline, and as result to a lower disease burden at the price of further socio-economic losses. Although it is very hard to assess the extent to which new restrictions would have been put in place in case of an epidemic resurgence, in the Supplementary Information, we investigate a scenario where NPIs as strict as those put in place in the second pandemic wave would have been used to contrast the disease resurgence in absence of vaccines. In other words, we modified the baseline against which the impact of vaccines is measured. The results indicate that, even with a very strong reduction of socio-economic activities, the absence of vaccines would have led to 12*K*+ more deaths and 540*K*+ infections. These numbers are in line with estimates recently reported in Ref. [19].

### Counterfactual vaccination scenarios

We implement several counterfactual scenarios to assess the effectiveness and quantify the impact of different vaccines allocation strategies. Along with the real vaccine allocation, that we will call *actual strategy*, we consider two additional counterfactual strategies. In *strategy 2* we imagine a scenario where vaccines are allocated in decreasing age order starting from the 80+. This allocation strategy aims at reducing disease severity by targeting first individuals at higher risk of facing severe outcomes such as hospitalization and death. It is important to mention that the *actual strategy* and *strategy 2* are very similar. However, in the actual rollout strategy some categories, for example teachers and other professional category, were added to the list of priority (in some regions) thus not respecting the strict age order we consider in *strategy 2*. In *strategy 3*, instead, we first allocate vaccines to the age groups 20 − 49 and then homogeneously to the rest of the population. This strategy aims at reducing disease transmission prioritizing individuals that are socially active. It is important to notice how both strategies account for front-line workers and the fragile population as recorded in the data. For more details on the vaccine allocation scenarios see the Materials and Methods section.

In Fig. 4A we compare these strategies in terms of averted COVID-19 deaths and infections. Since the different regions considered have different populations, we express averted deaths and infections as percentages with respect to the baseline simulations without vaccines. We see a common pattern emerging. Indeed, in all regions *strategy 2* is the most effective in reducing the number of deaths, followed by the *actual strategy* and *strategy 3*. As a concrete example, we estimate that in South Italy vaccines averted 29% (*IQR*: [19%−37%]) of the deaths that would have been observed without vaccines. This figure increases to 32% (*IQR*: [22% − 40%]) when the *strategy 2* is considered, while it drops to 20% (*IQR*: [9% − 30%]) with *strategy 3*. When instead averted infections are considered, we find the ordering inverted: *strategy 3* is the most efficient in reducing COVID-19 infections, followed by the *actual strategy* and *strategy 2*. This is in line with previous findings in the context of COVID-19 vaccination modeling [34, 35]. At the national level, we estimate that the strategy targeting first strictly the elderly would have prevented more deaths (31, 786, *IQR*: [19, 115−44, 733]) with respect to the *actual strategy* (29, 350, *IQR*: [16, 454−42, 826]), while a strategy prioritizing the younger would have avoided much less deaths (21, 440, *IQR*: [8, 006 − 35, 429]). Similarly, we compare strategies according to their impact on the Infection Fatality Rate (IFR), defined as the fraction of infections that result in death. In Fig. 4B we show, at both regional and national level, the percentage reduction of the IFR achieved as of 2021/07/05 with different vaccine allocation strategies (medians and IQRs are reported). We note a much more marked distinction between strategies with respect to results in Fig. 4A. This is not surprising, indeed the same reduction in deaths, for example, can be achieved both reducing mortality or the number of people that are reached by the disease. As expected, across the different regions, when vaccines are not considered the IFR remains constant (i.e., reduction of − 0.06%, *IQR*: [− 0.99%;1.27%]). The *strategy 2* is instead the most effective one in reducing the IFR, followed by the *actual strategy*, and finally *strategy 3*. Indeed, we estimate that in Italy vaccines reduced COVID-19 IFR by − 22.2% (*IQR*: [− 31.4%; − 13.9%]), while a strategy prioritizing strictly the elderly would have implied a reduction of − 29.2% (*IQR*: [− 38.2%; − 21.2%]). On the other hand, a strategy targeting the younger would have had a very small impact on IFR (− 2.1%, *IQR*: [− 6.0%; 1.5%]).

**Figure 4:**
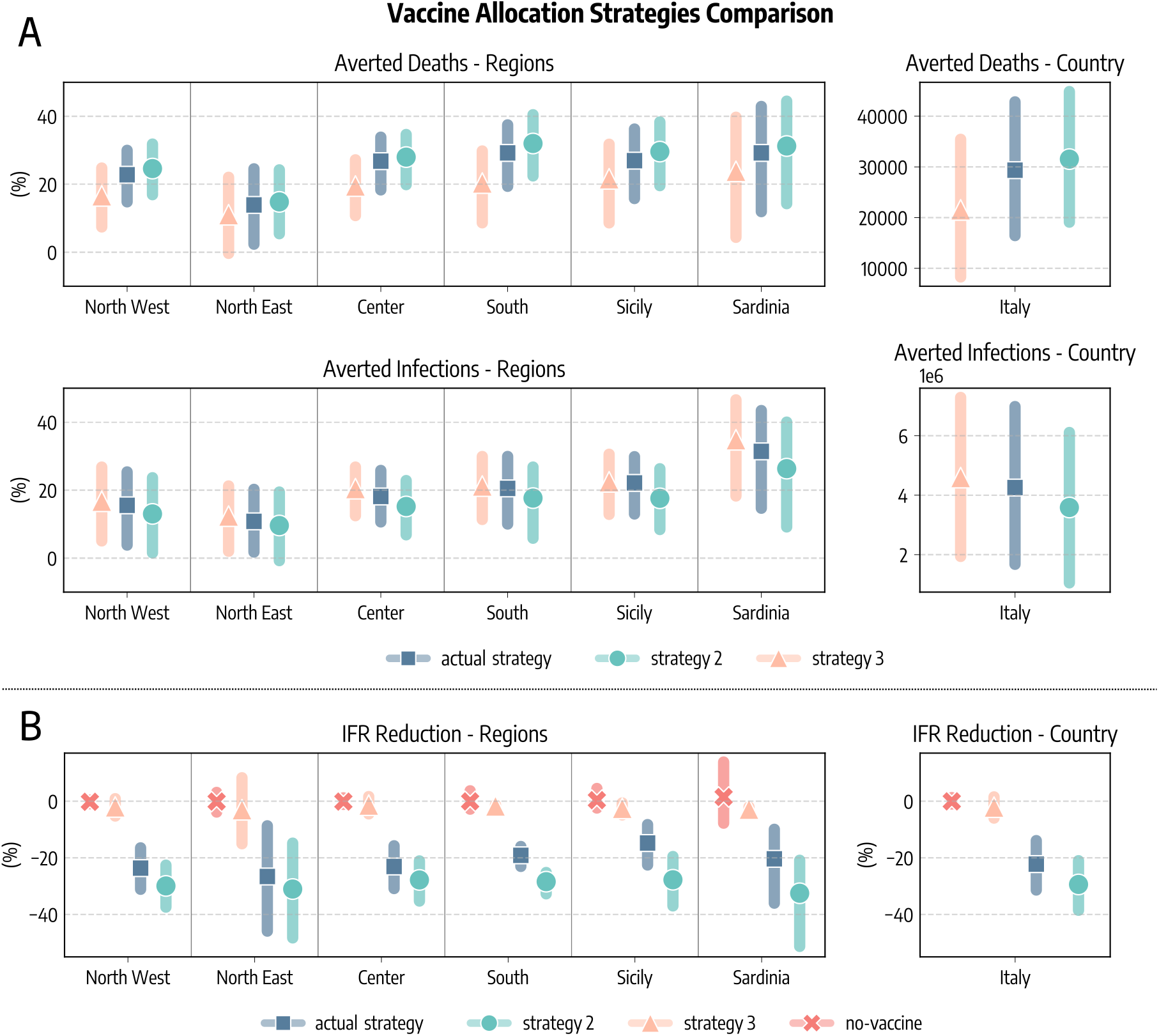
Comparison of vaccine allocation strategies. A) Averted COVID-19 deaths and infections, both at subnational and national level (medians and interquartile ranges are reported). B) Percentage reduction of the Infection Fatality Rate as of 2021/07/05. In all panels, *actual strategy* denotes an allocation strategy that follows the observed allocation as it unfolded during the pandemic, *strategy 2* considers the case where vaccines are allocated in decreasing age order starting from the 80+, *strategy 3* considers the case in which vaccines are first allocated to the age groups 20 − 49 and then homogeneously to the rest of the population, and *no-vaccine* denotes the counterfactual scenario in which vaccines were not administered.

One additional question concern what could have happened in Italy with the availability and timeline of vaccination programs as in other countries such as United Kingdom and the United States. In Fig. 5 we show averted deaths considering the vaccine allocation and rescaling doses to match vaccination rates in the UK and US. At the sub-national level, averted deaths (median and IQR) are expressed as percentage of fatalities observed in the simulations without vaccines. We also report the total number of averted deaths (median and IQR) at the country level. We estimate that 46, 046 (*IQR*: [33, 696−59, 022]) deaths would have been averted if Italy had the same availability of vaccines of the UK, and 42, 578 (*IQR*: [29, 768 − 55, 835]) when matching the availability of the US. This implies approximately an additional 16, 700 and 13, 200 lives saved with respect to the actual rollout case in the case of, respectively, UK and US. We note how the total number of doses administered in the period under study differs in the three scenarios. We see in Fig. 5 that as of 2021/07/05 33.1*M* doses are administered in Italy, 41.8*M* when matching the UK, and 35.4*M* when matching US.

**Figure 5:**
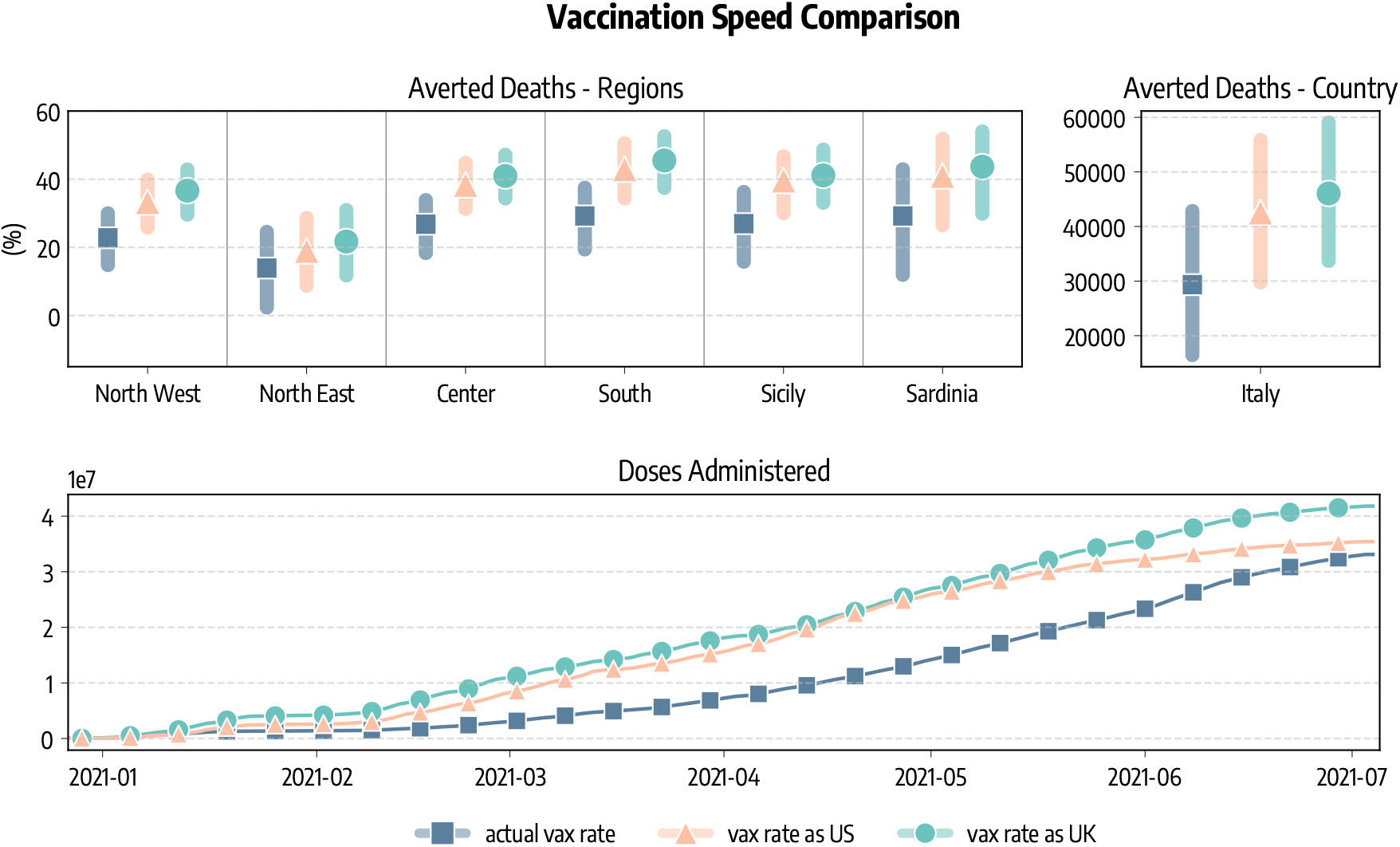
Comparison of vaccination speeds and doses. We show the percentage of deaths averted with respect to simulations without vaccines in different regions considering the real data-driven campaign, and rescaling the number of doses to match those administered in the United Kingdom and the United States. We also report the estimated total number of deaths (median and IQR) averted in Italy as well as the number of cumulative doses administered in the scenarios.

## Discussion

We presented the *anatomy* of the first six months of the vaccine rollout in Italy. We showed that vaccines prevented an additional wave of infections after the partial reopening of the country in late April, 2021. These results (49 averted deaths per 100, 000) are in line with reports from the United Kingdom and the United States, where, respectively, in first six months of the rollout around 30^*′*^000 (45 per 100, 000) and 279, 000 (85 per 100, 000) additional deaths have been averted [14, 15].

Thanks to the mechanistic nature of our modeling approach, we analyzed and compared different vaccines allocation strategies through counterfactual scenarios. The strategy strictly prioritizing the elderly would have prevented more deaths and contributed to a higher reduction in the observed IFR. On the other hand, prioritizing the most active age groups (i.e., younger) we would have averted more infections but lead to more fatalities. Similarly, rescaling the number of available doses to those delivered in other countries of reference, we tested the effects of faster vaccination rates.

The present work comes with limitations. First, the compartmental setup used to model disease progression is a relatively simple one compared to other approaches that consider, for example, also the pre-symptomatic and asymptomatic stages of the infection [36, 37]. Nonetheless, it has been previously used in several works in the context of COVID-19 modeling [22, 38–40]. Second, both the vaccination protocol and the effect of vaccines on disease progression are an approximation of reality [41]. For simplicity we considered, besides the wild type, only one additional virus strain, although we acknowledge that the Alpha variant was not the only variant of concern circulating in Italy during the period considered [42]. Beside the importation data from GLEAM, we model each region separately, thus neglecting the coupling between them via different forms of mobility. Finally, in the counterfactual scenarios we considered all individuals willing to receive vaccines. While the current vaccination rates in Italy show a high vaccine acceptance (81.5% of the population 12+ completed the vaccination course and 85.7% received at least one dose as of 2021/10/19 according to official sources [43]), this is an optimistic assumption. In the Supplementary Information we relax this and we study the effect of vaccine hesitancy. We measured the effects of vaccines respect to a baseline that considers the observed contact patterns during the rollout. As mentioned above, the resurgence of infections and deaths, that would have been observed without vaccines, would have led to new NPIs aimed at limiting the chains of infections. In the Supplementary Information, we investigate a different baseline where an uptick of cases and deaths is associated to NPIs as strict as those put in place in second pandemic wave (i.e. fall 2020). Interestingly, even with a very strong reduction of socio-economic activities the lack of vaccines would have led to 12*K*+ extra deaths and 540*K*+ infections. These number are in line with the recent estimates presented in Ref. [19]. Hence, it is important to acknowledge how the selection of the baseline against which the impact of the rollout is quantified affects the results. As we have experienced throughout 2020, very restrictive NPIs can reduce the burden of the disease in absence of vaccines. However, it is very hard to estimate what would have been the response to a new pandemic wave induced by the Alpha variant and the lack of vaccines, especially considering the NPIs fatigue of the population after months of restrictions.

In conclusion, we combined mathematical modeling and data to provide a realistic representation of the interplay of COVID-19 spread, vaccines rollout, NPIs, and the emergence of a more transmissible virus strain. The results highlight the strong positive impact and key role of vaccines in the evolution of the COVID-19 pandemic in Italy. While we have focused only on the Italian context, our approach can be easily extended to other countries helping to characterize and evaluate vaccination campaigns worldwide.

## Materials and Methods

### The epidemic model

Individuals who are susceptible to the disease are placed in the *S* compartment. Interacting with the infectious, they can get infected and transition to the Latent *L* compartment. After the latent period *ϵ*^−1^, *L* become infectious and enter the compartment *I*. Lastly, after the infectious period *µ*^−1^, *I* individuals transition to the Removed compartment *R* (a schematic representation of the compartmental structure is provided in Fig. 6). Similar approaches have been previously used to model disease progression in the context of COVID-19 [22, 39, 40]. Furthermore, we account for the age-stratification of the population and of their contacts. Individuals are divided into the following 10 age groups: 0−9, 10−19, 20−24, 25−29, 30−39, 40−49, 50−59, 60−69, 70−79, 80+. The number of contacts between age groups is defined by the country-specific contacts matrix *C* from Ref. [26]. We simulate the number of daily deaths considering removed individuals for each age group and the age stratified Infection Fatality Rate (IFR) from Ref. [44]. To account for delays between the transition *I* → *R* and actual death, we record fatalities computed on the recovered of a certain day only after Δ days. In addition, we introduce a seasonal modulation to model variations in factors such as humidity and temperature that can influence transmissibility [23, 27]. In practice this implies a rescaling of the effective reproductive number *R*_*t*_ → *s*_*i*_(*t*)*R*_*t*_, with *s*_*i*_(*t*) equal to the following function:

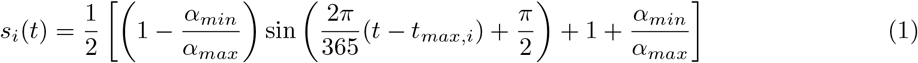

**Figure 6:**
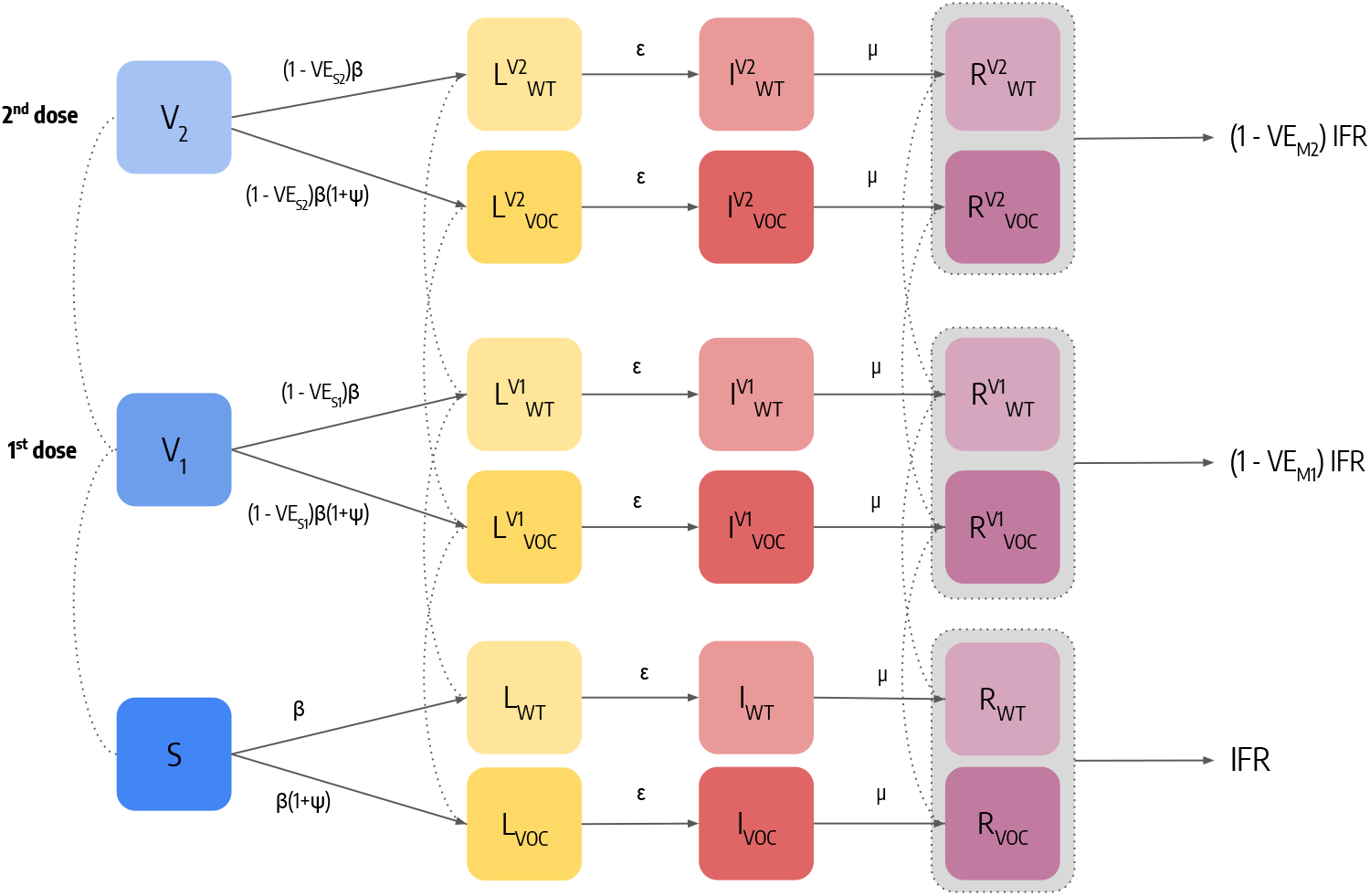
Schematic representation of the epidemic model and transitions between compartments. For simplicity, we represent the model for a single age group.

Where *i* refers to the hemisphere considered, and *t*_*max,i*_ is the time corresponding to the maximum of the sinusoidal function. For the northern hemisphere it is fixed to January 15^*th*^. We set *α*_*max*_ = 1 and consider *α*_*min*_ as a free parameter (see more details below).

Additional Latent and Infectious compartments are included to model a second, more transmissible virus strain. Given *β* as the transmission rate of the wild type, the second strain has a rate *β*(1 + *ψ*), where *ψ* captures the increased transmissibility. We assume that the this second strain has the same latent and infectious period as well as IFR of the wild type, and we set *ψ* = 0.5, compatible with the characteristics of the Alpha variant [7, 10]. In the Supplementary Information we repeat the analyses considering different values of *ψ* and an increased IFR. Individuals infected with the second strain are initialized considering realistic estimates of Alpha variant importations from GLEAM [22, 23, 31] during the period 2020/09-2020/12, more details on importations are provided below.

**Table 1:** Vaccine efficacy.

| | $VE_1$ ($VE_{S1}$) | | $VE_2$ ($VE_{S2}$) | | $\Delta_{V2}$ |
| --- | --- | --- | --- | --- | --- |
|  | Wild Type | Alpha | Wild Type | Alpha |  |
| *Pfizer* | 90% (80%) | 50% (40%) | 95% (90%) | 90% (80%) | 21 days |
| *Moderna* | 90% (80%) | 50% (40%) | 95% (90%) | 90% (80%) | 28 days |
| *AstraZeneca* | 70% (60%) | 50% (40%) | 80% (70%) | 75% (65%) | 90 days |
| *Janssen* | 70% (60%) | 50% (40%) | / | / | / |

Finally, we include specific compartments to model the vaccination campaign. Our general modeling setup accommodates both single and two doses vaccines. Individuals who received one dose of vaccine move to the compartment *V*_1_ and see their probability of infection reduced by a factor 1 − *V E*_*S*1_, where *V E*_*S*1_ represents the effectiveness of vaccine against infection. If *V*_1_ individuals get infected, their IFR is also reduced by a factor 1 − *V E*_*M*1_. This implies that the overall efficacy of the 1^*st*^ dose is *V E*_1_ = 1 − (1 − *V* − *E*_*S*1_)(1 − *V E*_*M*1_). If the vaccine has a two doses regiment, *V*_1_ individuals then receive the second inoculation after Δ_*V* 2_ days and transition to the compartment *V*_2_. Similarly to the 1^*st*^ dose, the 2^*nd*^ dose provides an efficacy *V E*_*S*2_ and *V*_*M*2_ implying an overall *V E*_2_ = 1−(1−*V E*_*S*2_)(1−*V E*_*M*2_). We also consider that all vaccinated individuals are less infectious by a factor (1 − *V E*_*I*_) (*V E*_*I*_ = 40% [45]) and that vaccines have reduced efficacy against the Alpha variant. We model separately the different vaccines authorized in Italy: Pfizer/BioNTech (Δ_*V* 2_ = 21*days*), Moderna (Δ_*V* 2_ = 28*days*), Vaxzevria/AstraZeneca (Δ_*V* 2_ = 90*days*), an Janssen (single dose). Since vaccine protection is not immediate, we introduce a delay of Δ_*V*_ days between administration (of both 1^*st*^ and 2^*nd*^ dose) and actual effect of the vaccine. For example, an individual who received the 1^*st*^ dose on day *t*, will be protected with efficacy *V E*_1_ only after Δ_*V*_ days. Here we set Δ_*V*_ = 14*days*. Values of vaccine efficacy and Δ_*V* 2_ used in the simulations are reported in Tab. 1. We use Ref. [45] to inform the choice of different vaccine efficacy.

### Modeling of Non-pharmaceutical Interventions

The contacts matrix *C*, which defines the rates of contact between age groups, is made up of four contribution: contacts that happen at home (*C*^*home*^), school (*C*^*school*^), workplace (*C*^*work*^), and general community settings (*C*^*community*^). We model the variations in contacts induced by non-pharmaceutical interventions at workplaces and in the community settings using data from the COVID-19 Community Mobility Report published by Google [20]. More in detail, the report provides the positive or negative percentage variation *w*_*l*_(*t*) of number of visits to specific location *l* on day *t* with respect to a prepandemic baseline. We compute a proxy of contacts variation as follows: *r*_*l*_(*t*) = (1 − *w*_*l*_(*t*)*/*100)^2^. Indeed, the number of contacts between individuals is proportional to the square of their number. We use the field `workplaces percent change from baseline` to compute the contacts variations parameter in workplaces, and the average of the fields `retail and recreation percent change from baseline` and `transit stations percent change from baseline` for the general community settings. The data are provided at the level of the NUTS2 regions. We derive the contacts variation parameters for a NUTS1 region computing the weighted average with respect to the population of the parameters of the included NUTS2 territories. Lastly, we also perform temporal aggregation taking the weekly average of these parameters.

We model variations in contacts in schools using the timeline of policy interventions. More in detail, we reproduce the two phases of the Italian government approach to the implementation of containment measures. During the first one (before 2020/11/06), interventions where taken mostly at the national level. We capture the effects of NPIs on schools in this phase using data from the Oxford COVID-19 Government Response Tracker [21]. The report provide daily indexes expressing the strictness of policies regarding school closures at the national level. During the second phase, regions where divided into risk zones according to the local state of the epidemic [46]. Each risk zone had specific rules regarding schools and allowed activities. We model this taking into account the actual time-varying definition of risk zones in Italy. Full details are provided in the Supplementary Information.

### Modeling Introductions of the Alpha Variant

We model the introduction of Alpha variant infections in each geographical area using GLEAM, a global stochastic metapopulation model that simulates the mobility of people across more than 3,300 sub-populations in about 190 countries/territories [22, 23, 31]. Sub-populations are defined by the catchment area of major transportation hubs and mobility among them includes both long-range air travel (obtained from the International Air Transport Association and Official Airline Guide (OAG) databases) and short-scale commuting patterns. Origin-destination data on passengers provided by the OAG [47] from 2020 are used to model international airline travel. The model is calibrated to importation of cases from China at the beginning of the Pandemic as well as the evolution of deaths in each country. GLEAM accounts for travel limitations, mobility reductions, and government interventions. We account for the stochastic nature of importations and onset of local transmission considering 307, 000 stochastic simulations generated by the model. We consider arrivals of individuals in the latent compartment only for each age bracket. Indeed, travelers were required to show a negative test and other measures were implemented in airports to prevent symptomatic individuals to travel. The first two specimens of the Alpha variant were collected on September 20 and 21, 2020 in London and in the Kent area. As the UK sequences about 5% of positive cases [48], we modeled the emergence of the Alpha variant on week 38 of 2020 assuming a cluster of symptomatic/exposed infectious individuals drawn from a Poisson distribution with mean value of 40 symptomatic individuals. In the main text, we assume the new variant as 50% more transmissible (i.e., *ψ* = 0.5), in line with current estimates [7].

### Counterfactual Vaccine Allocation Scenarios

We investigate the impact of two counterfactual vaccination strategies in which we change the allocation strategy. *Strategy 2* aims at mitigating the spread by reducing the severity of the disease. This is achieved prioritising the part of population exposed to higher risks of facing severe outcomes (e.g., death) if infected. Since the IFR of COVID-19 strongly correlates with age, in this scenario we start vaccinating the age group 80+ and then we proceed in strictly decreasing order of age until all 50+ individuals are vaccinated. After, we distribute vaccines homogeneously to the population under 50 since we assume that vaccines are made available to everyone after that individuals associated with higher IFR have been vaccinated. With *strategy 3* disease mitigation is achieved by reducing transmission rather than severity. In our simulations, this translates in targeting first the most active age groups and then the rest of the population. Following previous work in the context of COVID-19 vaccination campaign, we select the age groups 20 − 49 as the primary target of this allocation strategy [34, 35]. These former studies also showed that prioritising the elderly is preferable in terms of number of averted deaths, while vaccinating the younger is more efficient at reducing cumulative incidence. In both scenarios, we still give vaccines according to the real data to healthcare workers, care facilities residents, and people with comorbidities that are considered risk factor for COVID-19. Official sources stopped to provide the information regarding the categories of the vaccinated individuals on 2021/05/26, therefore after this date we allocate vaccines following only the strategy considered. Vaccines in Italy were administered to the 12+ population. To match our age group resolution, in the counterfactual vaccine allocation strategy 2 and 3 we only give vaccines to the 10+ individuals. In Fig. 7 we show the average age of vaccinated individuals in different allocation strategies. Across the regions considered, we observe that at the beginning of the rollout different strategies look similar. Indeed, in that period received the vaccines healthcare workers and fragile individuals that are accounted for also in the counterfactual scenarios as mentioned previously. Since the beginning of February 2020, instead, strategies start to differ. In particular, the strategy targeting the elderly (*strategy 2*) shows a higher average age with respect to the (*actual strategy*). This is because, despite elder individuals were prioritized in the actual vaccination campaign in Italy, also other categories were initially given a priority, such as teachers. Reasonably, the strategy targeting first younger individuals (*strategy 3*) shows a lower average age of vaccinated with respect to the other two. In the counterfactual strategies we assume that all individuals are willing to receive the vaccine.

**Figure 7:**
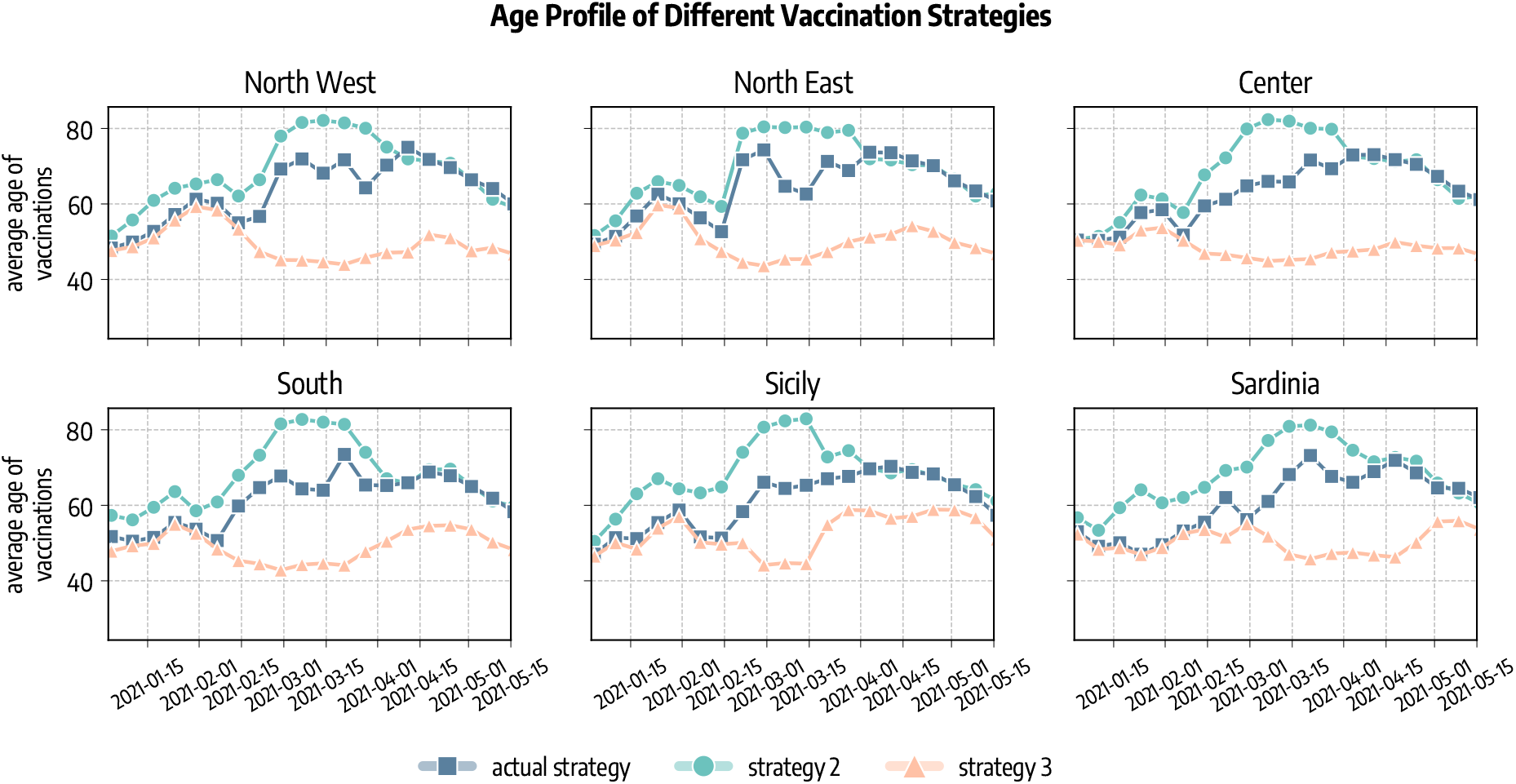
Average age of vaccinated in different allocation strategies. *Actual strategy* denotes the observed vaccine allocation as it unfolded during the pandemic, *strategy 2* considers the case where vaccines are allocated in decreasing age order starting from the 80+, and *strategy 3* considers the case in which vaccines are first allocated to the age groups 20−49 and then homogeneously to the rest of the population.

We consider additional counterfactual scenarios in which we apply to Italy the vaccination rates of other countries of reference, namely United Kingdom, and United States. These countries administered more vaccines than Italy as of 2021/07/05, and were faster especially during the early months of the rollout. We rescale the number of doses as follows. If in Italy on day *t* were administered *x*_*t*_ doses per person (*X*_*t*_ in total), while in the other country *y*_*t*_, in the counterfactual scenario we deliver on day *t* a number of doses 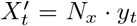 (where *N*_*x*_ is the Italian population). We stress how we only change the number of available doses while we keep, in all scenarios, the same data-driven age allocation strategy and the same vaccines administered. Indeed, we do not aim to replicate exactly the vaccination campaigns of other countries of reference, but only to test different rollout rates.

### Model Calibration

We calibrate the free parameters of the model using an Approximate Bayesian Computation (ABC) technique [25, 30]. We define the prior distributions of the free parameters *P*(*θ*), a number accepted sets *N*, an error metric *m*(*E, E*′), and a tolerance *δ*. We start sampling a set of parameters *θ* from *P*(*θ*) and generate an instance of the model using these parameters. Then, using the chosen error metric we compare an output quantity *E*′ of the model with the corresponding real quantity *E*: if *m*(*E, E*′) *< δ* then we accept the set *θ*, otherwise we reject it. We repeat this accept/reject step until *N* parameter sets are accepted. The empirical distribution of the accepted sets is an approximation of their real posterior distribution. We produce model’s projections sampling parameters sets from the pool of accepted sets and generating an ensemble of possible epidemic trajectories. We then compute median and confidence intervals on this ensemble. In this work, we consider the following free parameters and priors:

**Table 2:**
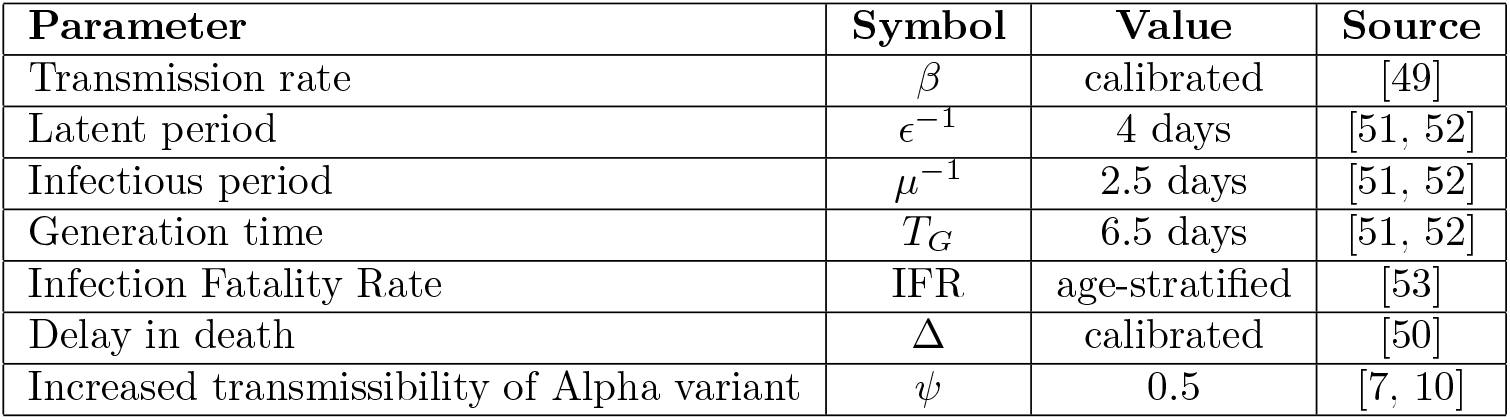
List of parameters and related sources.

- *β*, we explore values of the transmission rate parameter such that the associated 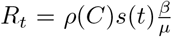 on the first simulation day ∼ *U* (1.2, 2.0) [49];
- Δ ∼ *U*(14, 25). The calibration range is informed by Ref. [50];
- *α*_*min*_ ∼ *U*(0.5, 1.0), in doing so we explore values from strong (0.5) to absent (1.0) seasonality;
- we select initial conditions from an ensemble of realistic estimates from GLEAM. Each estimate has the number of individuals of different age groups in different compartments (*S, L, I, R*) on the start of the simulation (2020/09/01).

The model is calibrated separately for each of the six regions considered. We set the calibration period to 2020/09/01-2021/07/05. We consider weekly deaths as output quantity and the weighted mean absolute percentage error (wMAPE) as error metric. We also set the number of accepted sets *N* = 3000 and the tolerance *δ* = 0.40 for Sicily and Sardinia and *δ* = 0.35 for the remaining regions. For the different basins, we represent in Fig. 8 the number of weekly real and simulated deaths (median and 90% CI). We obtain a median wMAPE of 0.23 for North West, 0.26 for North East, 0.25 for Center, 0.18 for South, 0.30 for Sicily, and of 0.40 for Sardinia. In the Supplementary Information we report the posterior distributions of the free parameters obtained through the ABC calibration.

**Figure 8:**
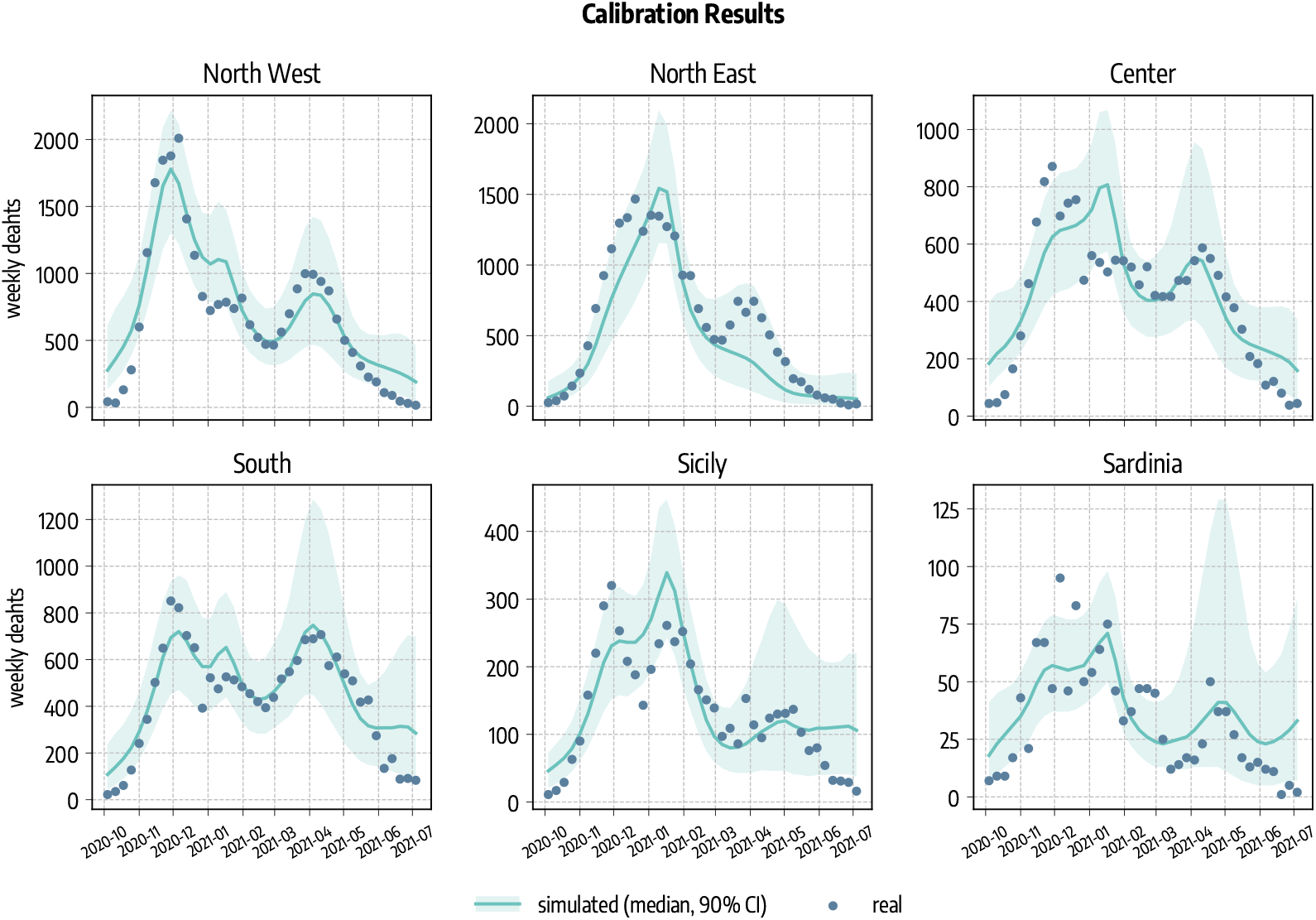
Calibration Results. Simulated (median and 90% CI) and observed cumulative number of deaths.

## Data Availability

All the data used in the paper are publicly available and the sources are cited within the text.
The data produced in the study are available upon request to the authors

## Acknowledgements

All authors thank the High Performance Computing facilities at Greenwich University. N.G. acknowledges support from the Doctoral Training Alliance. A.V. acknowledges support from NIH-R56AI148284 award. M.A. and A.V. acknowledge support from the Bill Melinda Gates Foundation (award number INV006010) and the Google Cloud Research Credits program to fund this project. The findings and conclusions in this study are those of the authors and do not necessarily represent the official position of the funding agencies, the National Institutes of Health, or the U.S. Department of Health and Human Services.

## Author Contributions

N.G, M.C, N.P. and A.V designed the research. N.G. performed the simulations. N.G., M.C, N.P. and A.V. wrote the first draft of the manuscript. All authors contributed interpreting the data, editing and approving the manuscript.

