## Supplementary Information for "Anatomy of the first six months of COVID-19 Vaccination Campaign in Italy"

### Supplementary Material for *Anatomy of the first six months of COVID-19 Vaccination Campaign in Italy*

#### Contents

**1 Posterior Distributions 2**

**2 Modeling non-pharmaceutical interventions: school closures 2**

**3 Vaccine Hesitancy 4**

**4 Different Transmissibility of Alpha Variant 5**

**5 Increased Mortality for the Alpha Variant 7**

**6 The Role of Asymptomatic and Recovered on Vaccinations 8**

**7 Baseline with behavioral reactions 8**

### 1 Posterior Distributions

In Fig. 1 are shown the posterior distributions of the parameters calibrated through the ABC rejection algorithm [1, 2] for each region. In particular, we report the effective reproductive number $R_t$ on the first simulation day, the delay (expressed in days) in deaths $\Delta$, the seasonality parameter $\alpha_{min}$, and the initial number of infected ($I$ plus $L$ individuals) and recovered per $100'000$ at the start of the simulation.

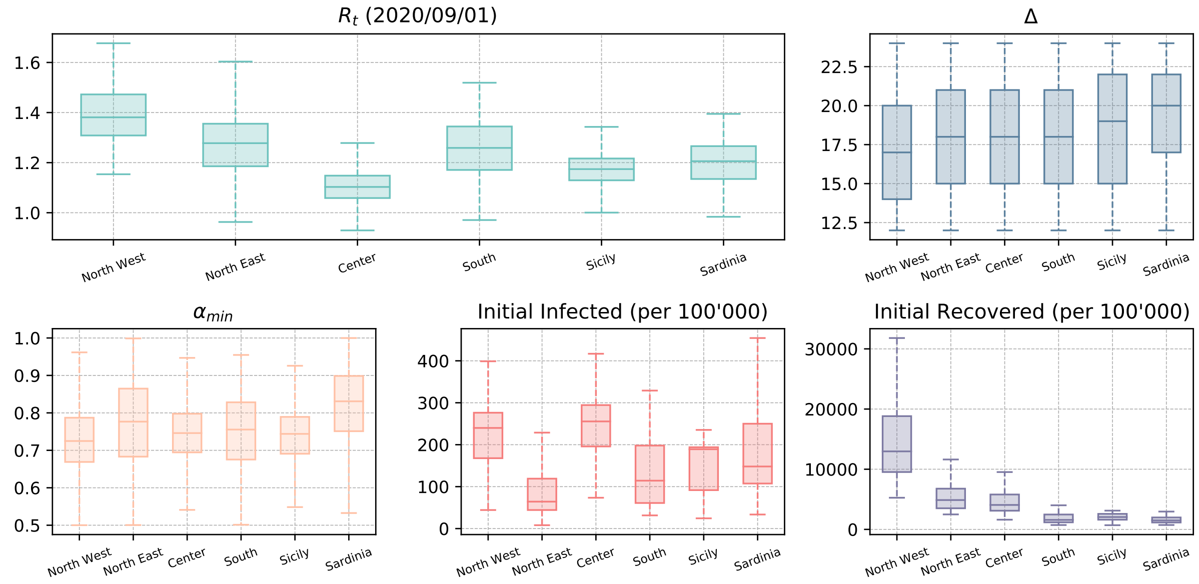


Figure 1: **Posterior distributions of parameters obtained through ABC calibration**. Each boxplot summarises the distribution of the parameter showing median, interquartile range (IQR), and *minimum* and *maximum* values defined respectively as $Q1 - 1.5IQR$ and $Q3 + 1.5IQR$ (where $Q1$ and $Q3$ are the first and third quartile).

### 2 Modeling non-pharmaceutical interventions: school closures

We model variations in contacts in schools as follows. First, we identify two phases. Before 2020/11/06, interventions from the Italian Government were mostly at the national level and different regions followed the same rules and recommendations. During this first phase, we use data from the Oxford COVID-19 Government Response Tracker [3] to model contacts variation in schools. We consider three indexes from this report about school closure ($school_{idx}$), workplace closure ($work_{idx}$), and stay at home requirements ($stayhome_{idx}$). These indexes vary daily and are provided at the national level. They can assume discrete values from 0 (no measures) to 3 (maximum restrictions). We define the parameters $w_s(t)$ and $\alpha(t)$ as follows:

1. $school_{idx}(t) = 0 \rightarrow w_s(t) = 1.0,\ \alpha(t) = 1.0$
2. $school_{idx}(t) = 1 \rightarrow w_s(t) = 0.5,\ \alpha(t) = 1.0$
3. $school_{idx}(t) = 2 \rightarrow w_s(t) = 1.0,\ \alpha(t) = 0.3444$
4. $school_{idx}(t) = 3$ and NOT$(work_{idx}(t) \geq 2$ and $stayhome_{idx}(t) \geq 2) \rightarrow w_s(t) = 1.0,\ \alpha(t) = 0.3444$
5. $school_{idx}(t) = 3$ and $(work_{idx}(t) \geq 2$ and $stayhome_{idx}(t) \geq 2) \rightarrow w_s(t) = 0.0,\ \alpha(t) = 1.0$

Finally, we model the impact of school closing on contacts on day $t$ multiplying the school layer $C^{school}$ (the elements describing contacts between $0-24$ years old individuals of the overall contacts matrix $C$) by $w_s(t)$ and by $\alpha(t)$. The two indexes are introduced to modify contacts that the younger age groups may have at school but also in other settings. The idea behind our approach is to define increasingly higher risk levels according to the policies implemented. The first case, for example, refers to a situation with no restrictions in place, therefore contacts in schools are not affected. When instead $school_{idx}(t) = 1$ some restrictions are in place, school closing or hybrid teaching mode and additional NPIs in classroom are recommended resulting in significant differences compared to normal operations [3]. We assume this would result in significant reduction of contacts in school. When $school_{idx}(t) = 2$ we introduce a reduction factor on the contacts of $0-24$ which is taken from Ref. [4], where authors studied the impact of school closure on contacts patterns among younger population. In the fourth case, strict closure policies on school are in place ($school_{idx}(t) = 3$) but younger age groups may still meet in different context apart from school (i.e., the work from home and the stay at home indexes are not at the maximum value). We assume that this would result in the same contacts reduction of the previous cases. Finally, when closure is required at all levels we send to zero the layer of contacts in schools.

After 2020/11/06, the Italian government introduced a tier system in which different NUTS2 regions were assigned to one of four risk levels (from 0, lowest risk, to 4 highest risk) [5]. Different tiers are associated with increasingly more restrictive measures. Because of our spatial resolution, first we define the risk tier at the NUTS1 level as the weighted average (according to the population) of the risk levels of the component NUTS2 regions (rounded to the nearest integer). For example, for NUTS1 region $R$, the risk tier at time $t$ is:

$$TIER_R(t) = \left\lfloor \frac{\sum_{r \in R} p_r * TIER_r(t)}{\sum_{r \in R} p_r} \right\rceil \qquad (1)$$

Where the summation runs over all NUTS2 regions $r$ which are part of NUTS1 region $R$, and $p_r$ is the population of NUTS2 region $r$ taken from official sources [6]. Finally, we define the parameters $w_s(t)$ and $\alpha(t)$ similarly to what we did before for the first phase:

1. $TIER_R(t) = 0 \rightarrow w_s(t) = 0.5,\ \alpha(t) = 1.0$
2. $TIER_R(t) = 1 \rightarrow w_s(t) = 0.5,\ \alpha(t) = 1.0$
3. $TIER_R(t) = 2 \rightarrow w_s(t) = 1.0,\ \alpha(t) = 0.3444$
4. $TIER_R(t) = 3 \rightarrow w_s(t) = 0.0,\ \alpha(t) = 1.0$

Then the contacts matrix for school on day $t$ is again obtained by the multiplying $C^{school}$ by $w_s(t)$ and by $\alpha(t)$ the contact rates between $0-24$ years old individuals.

#### 3 Vaccine Hesitancy

In the main text we assumed that in the counterfactual scenarios (*strategy 2* and *strategy 3*) all individuals are willing to receive the vaccines. Despite Italy showing very high rates of vaccine acceptance (as of 2021/10/19 85.7% received at least one dose [7]), this is an optimistic assumption. Therefore, we repeat here part of the analyses considering that a fraction of the population do not want to receive the vaccine. In Fig. 2 we show the the total number of averted deaths and infections (median and IQR) for *strategy 2* (prioritizing the elderly) and *3* (prioritizing the younger) for different values of hesitancy (i.e., the percentage of the population that refuses vaccines). We assume that hesitant individuals are homogeneously distributed among age groups. The horizontal dashed lines indicate the number of averted deaths/infections for the two strategies in the case of no hesitancy as presented in the main text. We observe, overall, a negligible effects of hesitancy. The explanation for such a small impact is that, as of 2021/07/05, *only* 58% of the population received at least one dose of vaccine in Italy. Therefore, the number of doses distributed in our simulations are not high enough to make the values of hesitancy considered significant. This also explains why we only observe an impact on model's outputs when considering averted deaths by *strategy 2*. In this scenario, we notice a decline in the number of averted deaths as the value of hesitancy grows. Indeed, since in this case we proceed in strictly decreasing age order between 50-80+, the values of hesitancy can play a role for older age groups that also feature higher IFRs.

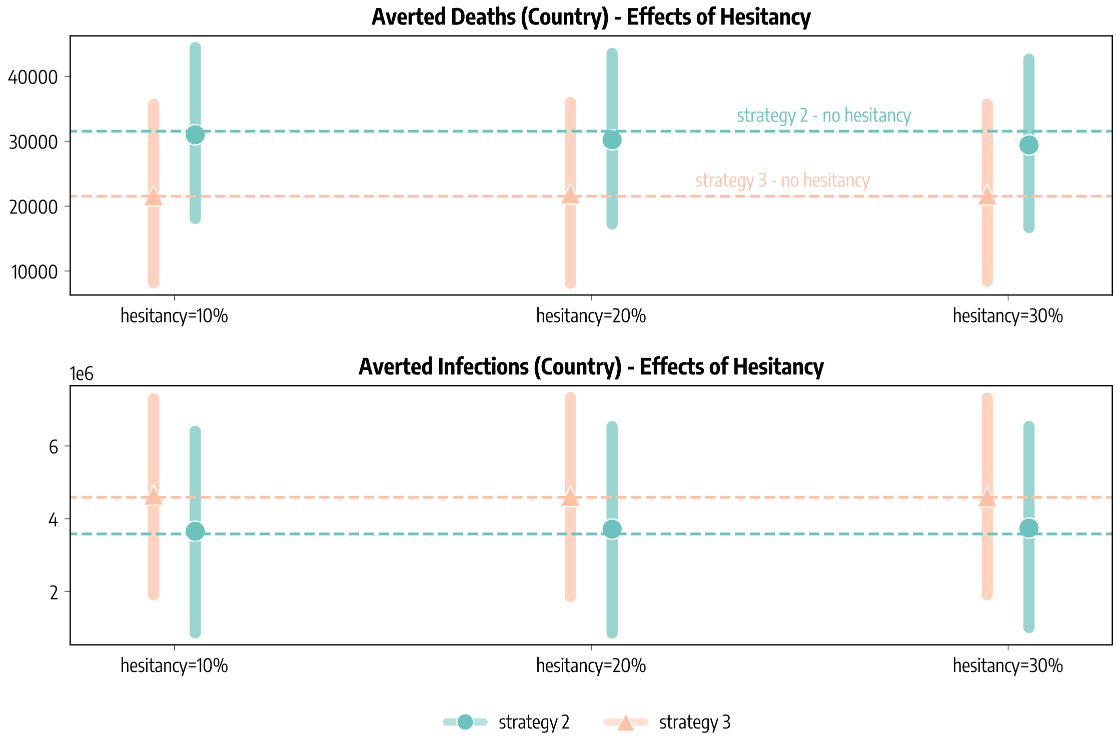


Figure 2: **Effects of vaccine hesitancy**

#### 4 Different Transmissibility of Alpha Variant

In the main text we assumed the variant Alpha to be 50% (i.e., $\psi = 0.5$) more infectious than the wild type circulating previously, in line with available estimates [8, 9]. Nonetheless, we provide a sensitivity analysis to the results considering different values of $\psi$. In particular, we consider here $\psi = 0.3$ and $\psi = 0.6$. We repeat the calibration step for the two additional values of $\psi$. We note a need to increase the ABC tolerance in the case of $\psi = 0.6$ for convergence reasons. In Tab. 1 we report the obtained wMAPE between simulated and reported weekly deaths for the three values of $\psi$. We notice how the calibration with $\psi = 0.5$ gives in general better results.

| | **wMAPE** | | |
|---|---|---|---|
| | $\psi = 0.3$ | $\psi = 0.5$ | $\psi = 0.6$ |
| North West | 0.33 | 0.23 | 0.28 |
| North East | 0.32 | 0.26 | 0.23 |
| Center | 0.32 | 0.25 | 0.32 |
| South | 0.28 | 0.18 | 0.27 |
| Sicily | 0.29 | 0.30 | 0.46 |
| Sardinia | 0.33 | 0.40 | 0.57 |

Table 1: **wMAPE between simulated and reported weekly deaths for different $\psi$**

In Fig. 3A we show averted deaths and infections at the subnational and national level. We observe how figures are lower when considering $\psi = 0.3$. This is expected. Indeed a less transmissible variant causes a milder wave in our simulations with no vaccine and therefore the difference with the simulations with vaccine administered is smaller. For the opposite reason, the number of averted deaths and infections is higher, instead, in the case of $\psi = 0.6$. At the national level, we estimate vaccines would have averted $12,896$ ($IQR$: $[7,002 - 19,254]$) deaths for $\psi = 0.3$ and $39,242$ ($IQR$: $[24,539 - 53,543]$) with $\psi = 0.6$. As discussed in the main for $\psi = 0.5$ we have instead $29,350$ ($IQR$: $[16,454 - 42,826]$). Similarly, the estimated number of averted infections would have been $2,383,158$ ($IQR$: $[748,270 - 4,130,513]$) with $\psi = 0.3$ and $4,815,666$ ($IQR$: $[2,411,182 - 7,207,004]$) with $\psi = 0.6$. For $\psi = 0.5$ we have instead $4,256,332$($IQR$: $[1,675,564 - 6,980,070]$).

In Fig. 3B we report the national prevalence of Alpha for the three values of $\psi$. When $\psi = 0.5$, we get a $wMAPE$ of 0.34 and a Pearson correlation coefficient $\rho = 0.84$ ($p < 0.001$) between the simulated prevalence and the one reported by genomic surveillance [10]. When $\psi = 0.3$ we get poorer results ($wMAPE = 0.53$, $\rho = 0.70$), and when $\psi = 0.6$ we get slightly better performance ($wMAPE = 0.31$, $\rho = 0.86$).

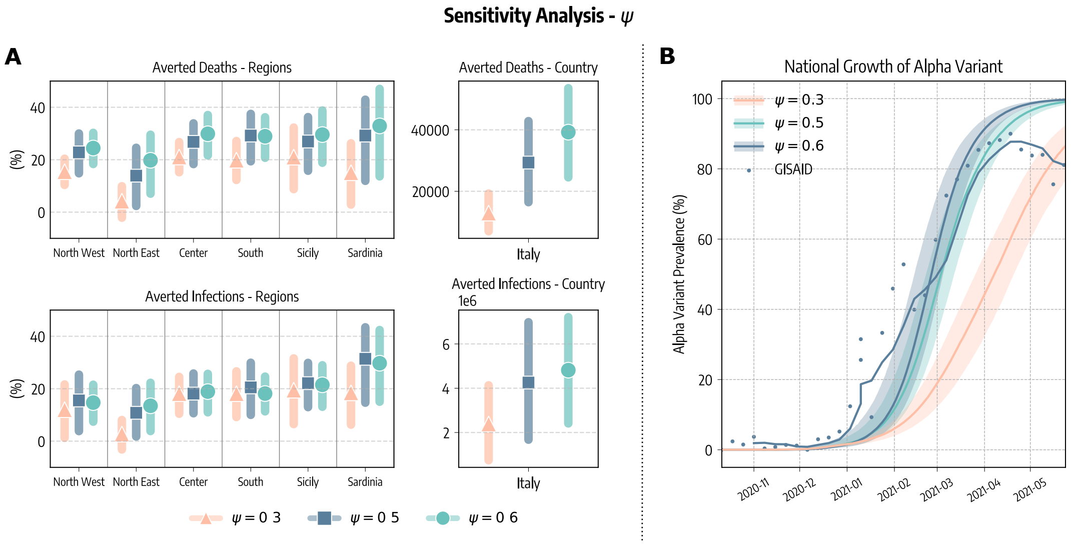


Figure 3: **Sensitivity analysis on $\psi$.** A) Averted deaths and infections at subnational and national level for different values of $\psi$. B) National prevalence of the Alpha variant as simulated with different $\psi$ and as reported by genomic surveillance.

#### 5 Increased Mortality for the Alpha Variant

In the main text we assumed the same Infection Fatality Ratio (IFR) for both the wild type and the Alpha variant. Nonetheless, previous works showed that the Alpha variant may also be associated with higher severity [11, 12]. To estimate how this impacts our findings we repeat here the analysis considering $IFR^{Alpha} = 1.5IFR^{wildtype}$, where $IFR^{wildtype}$ is the one used in the main text and is taken from Ref. [13], and the 1.5 factor is in line with previous works [11, 12].

In Fig. 4 we show averted deaths and infections using the same IFR for wild type and Alpha and using the higher IFR for Alpha. We observe that our findings are not significantly impacted by the change of the IFR. Overall, when the higher IFR for Alpha is considered we estimate a slightly higher number of averted deaths. For example, at the national level we estimate vaccines averted 29,350 ($IQR$: [16,454 − 42,826]) deaths when considering the same IFR for both strains, and 34,578 ($IQR$: [19,449 − 50,314]) when considering a higher IFR for Alpha.

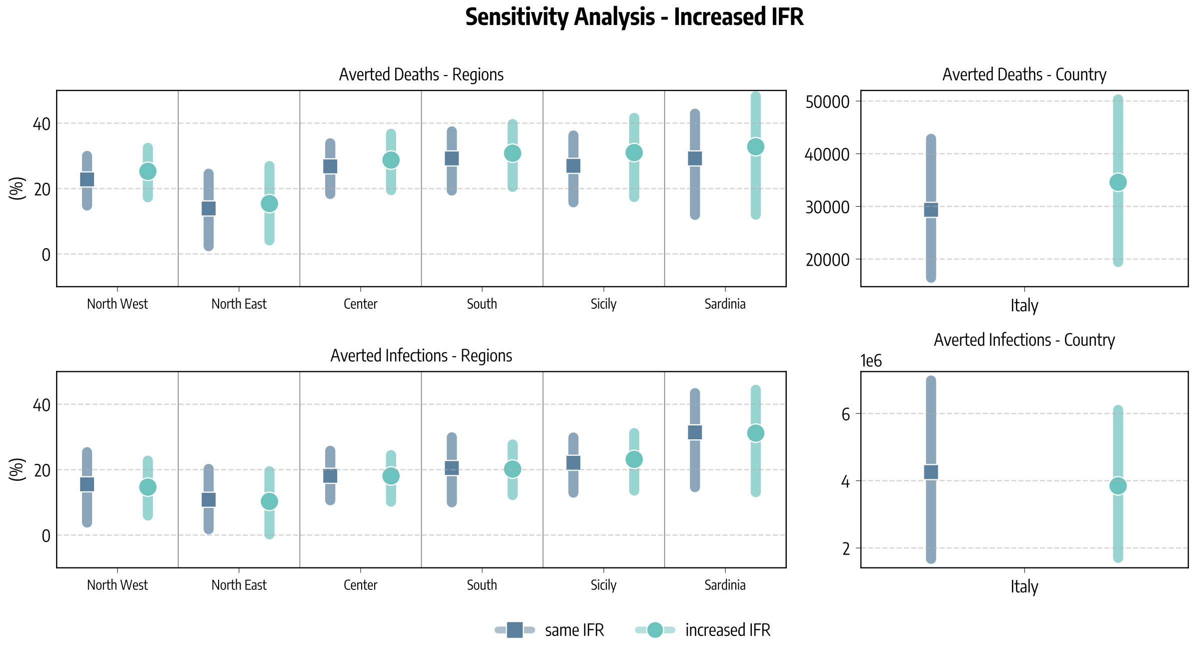


Figure 4: **Sensitivity analysis on Alpha variant IFR.** Averted deaths and infections at subnational and national level considering the same IFR for both wild type and Alpha and and a higher IFR for Alpha.

### 6 The Role of Asymptomatic and Recovered on Vaccinations

In the main text we assumed that all individuals but the infectious ($I$) can receive the vaccine. The real vaccination rollout in Italy, however, was more complex in terms of allocation. For example, individuals who recovered from the disease generally received only dose after at least three months from recovery and no more than six (then extended to twelve) [14]. At the same time, since vaccines were not officially preceded by a test, asymptomatic individuals may have received the vaccine as well. To assess how much the different allocation of vaccines among compartments can affect our result, we propose here the following analysis. In line with available estimates, we consider that 40% of all infections in our simulations are asymptomatic [15, 16]. Then, we assume that 40% of infectious individuals $I$ (i.e., the asymptomatic) can also receive the vaccines. We also assume that only 40% of recovered can receive the vaccine. Said differently, we assume that symptomatic recovered do not receive the vaccine. We note how this is an extreme scenario in which all asymptomatic infectious are eligible for vaccine and none of the symptomatic recovered are. The goal here is indeed to test the extent to which our results are impacted by different choices of allocation of vaccines among compartments.

In Fig. 5 we show averted deaths and infections using the allocation of vaccines among compartments presented in the main text and the one just described. We see that the change of allocation does not significantly impact our findings. For example, the number of averted deaths with the new allocation is $35,404$ ($IQR$: $[21,577 - 50,200]$), while in the main text we obtained $29,350$ ($IQR$: $[16,454 - 42,826]$). The higher number of averted deaths maybe be due to the fact that, with the new allocation among compartments, vaccines are given only to a fraction of the recovered and as result more doses are administered to susceptibles individuals.

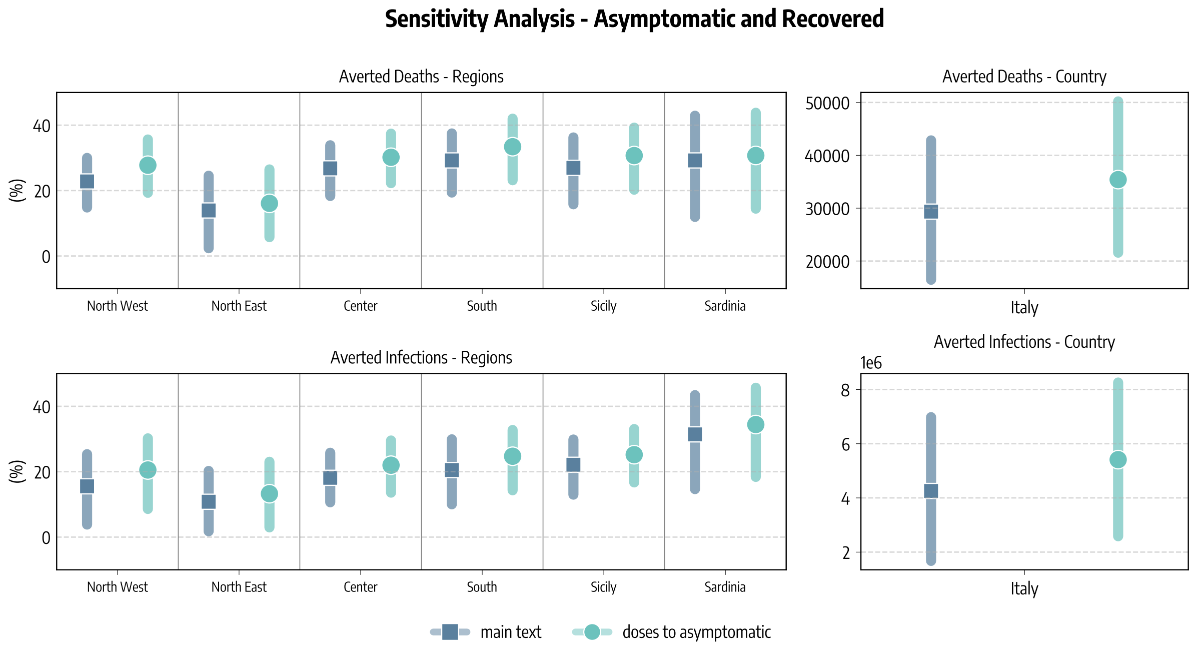


Figure 5: **Sensitivity analysis on allocation of vaccines among compartments.** Averted deaths and infections at subnational and national level considering the allocation among compartments proposed in the main text and considering also the role of asymptomatic and recovered.

### 7 Baseline with behavioral reactions

In the main text, the impact of vaccines is measured respect to a baseline that has everything equal but for the presence of vaccines. This approach allows us to isolate the impact of vaccines alone on the epidemic trajectory. Nonetheless, we acknowledge that the NPIs observed during the rollout were contingent to the improved epidemiological condition to which vaccines contributed in a substantial way. In the absence of vaccines, a worsening of the epidemiological conditions would have likely led to both bottom-up (i.e., self-initiated) and top-down (i.e., government mandated) behavioral reactions [17]. As result, the baseline is a pessimistic estimate since it does not account for possible NPIs enforcement in response to rises in infections and fatalities in case of no vaccinations.

Here, we present a second baseline without vaccines in which we also model behavioral reactions. It is important to stress that we will never know what it could have happened in the spring of 2021 without vaccines, so any scenario aiming to account for behavioral changes it is speculative. Bearing this in mind we proceed as follows. First, we define a threshold for the introduction of tougher restrictive measures. As mentioned in the main text, on 2020/11/06 the Italian government introduced a risk-level system to adopt more targeted and timely countermeasures according to the local epidemic conditions. Therefore, for each of our six regions, we compute the average number of deaths on the days when the region was moved to the highest risk level (indicated as ''*red zone*"). We call this quantity $D_i^{red\ zone}$. Second, we compute the average contacts matrix $C_i^{red\ zone}$ observed during the days when in region $i$ were in place the rules of the highest risk level. Finally, we follow this simple approach: in the simulation without vaccines administered, if the number of daily deaths in region $i$ exceeds $D_i^{red\ zone}$ - and the region is not already in the highest risk level - we use the contacts matrix $C_i^{red\ zone}$ instead of the one computed following actual mobility change and policy interventions. We apply this check after 2021/03/01 since it is around this date that the epidemic trajectories with and without vaccines start to visibly diverge.

We stress how this threshold approach to model NPIs enforcement has many limitations. For example, the system of risk-levels developed in Italy depended on many parameters and not just on deaths [5]. Also, we consider a single threshold, while in reality multiple thresholds associated with increasingly higher risk levels were defined. As result, our approach is admittedly simplistic to represent the complexity of individual and collective response to epidemics [18]. Nonetheless, it captures the possible evolution of NPIs in a hypothetical scenario using historical real data and allows us to assess the impact of this on our findings.

In Fig. 6 we compare the evolution of weekly deaths as reported by official sources and as simulated by our model. We consider the case in which i) vaccines are administered according to the real data, ii) no vaccines are administered, iii) no vaccines are administered but NPIs are put in place to counter the uptick of cases and deaths. As expected, in this last scenario we observe less deaths with respect to the simple baseline considered in the main. Nonetheless, it is important to stress how also in this case the death toll is worse with respect to the case with vaccines.

In Tab. 2 and Tab. 3 we compare the number of averted deaths and infections by vaccines with respect to the two no-vaccine baselines in different regions. Not surprisingly, we observe that total number of averted deaths with respect to the new baseline is diminished, from $29,4064$ ($IQR$: $[16,571; 42,830]$) to $12,676$ ($IQR$: $[2,795; 22,887]$). At the same time, we also obtain a much lower number of averted infections ($547,554$ $IQR$: $[-1,540,752; 2,701,933]$ respect to $4,306,913$ $IQR$: $[1,656,934; 6,990,557]$).

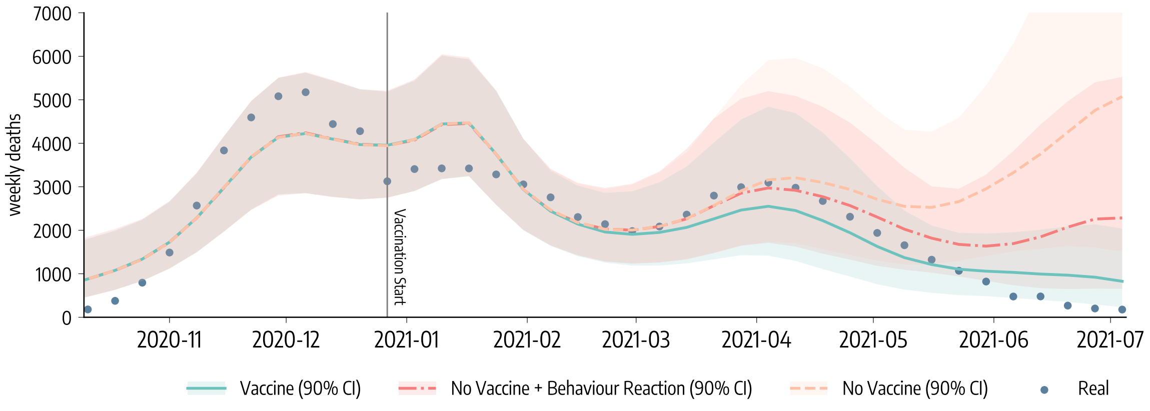


Figure 6: **Evolution of weekly deaths in Italy under different scenarios**. We show the number of weekly deaths in Italy as reported by official sources and s simulated by our model. We consider different vaccination scenarios: i) data-driven vaccination (solid line), ii) no vaccine (dashed line), and iii) no vaccine with possible behaviour reaction (dash-dotted line).

| Averted Deaths (Median and IQR) | | |
|---|---|---|
| | **No Vaccine Baseline** | **No Vaccine + Behavior Reaction Baseline** |
| *North West* | 9047 [5525, 12492] | 4672 [2052, 7480] |
| *North East* | 3143 [750, 5961] | 3128 [532, 5946] |
| *Center* | 6431 [4077, 8736] | 2469 [733, 4108] |
| *South* | 7892 [4745, 11235] | 1464 [-553, 3521] |
| *Sicily* | 2263 [1246, 3310] | 720 [56, 1387] |
| *Sardinia* | 629 [228, 1095] | 222 [-24, 444] |
| *Italy (total)* | 29406 [16571, 42830] | 12676 [2795, 22887] |

Table 2: **Averted deaths - No vaccine baseline comparison**

| Averted Infections (Median and IQR) | | |
|---|---|---|
| | **No Vaccine Baseline** | **No Vaccine + Behavior Reaction Baseline** |
| *North West* | 1011733 [269085, 1733792] | 260,281 [-390544, 886551] |
| *North East* | 506441 [43955, 993421] | 467160 [11869, 1008596] |
| *Center* | 875658 [460490, 1284045] | 75351 [-228365, 386458] |
| *South* | 1289381 [571882, 2044919] | -269856 [-745282, 201937] |
| *Sicily* | 451476 [243112, 652044] | 6086 [-134485, 155195] |
| *Sardinia* | 172223 [68409, 282335] | 8532 [-53945, 63195] |
| *Italy (total)* | 4306913 [1656934, 6990557] | 547554 [-1540752, 2701933] |

Table 3: **Averted infections - No vaccine baseline comparison**

[9] Erik Volz, Swapnil Mishra, Meera Chand, Jeffrey C Barrett, Robert Johnson, Lily Geidelberg, Wes R Hinsley, Daniel J Laydon, Gavin Dabrera, Áine O'Toole, Robert Amato, Manon Ragonnet-Cronin, Ian Harrison, Ben Jackson, Cristina V Ariani, Olivia Boyd, Nicholas J Loman, John T McCrone,

Sónia Gonçalves, David Jorgensen, Richard Myers, Verity Hill, David K Jackson, Katy Gaythorpe, Natalie Groves, John Sillitoe, Dominic P Kwiatkowski, Cherian Koshy, Amy Ash, Emma Wise, Nathan Moore, Matilde Mori, Nick Cortes, Jessica Lynch, Stephen Kidd, Derek J Fairley, Tanya Curran, James P McKenna, Helen Adams, Christophe Fraser, Tanya Golubchik, David Bonsall, Mohammed O Hassan-Ibrahim, Cassandra S Malone, Benjamin J Cogger, Michelle Wantoch, Nicola Reynolds, Ben Warne, Joshua Maksimovic, Karla Spellman, Kathryn McCluggage, Michaela John, Robert Beer, Safiah Afifi, Sian Morgan, Angela Marchbank, Anna Price, Christine Kitchen, Huw Gulliver, Ian Merrick, Joel Southgate, Martyn Guest, Robert Munn, Trudy Workman, Thomas R Connor, William Fuller, Catherine Bresner, Luke B Snell, Amita Patel, Themoula Charalampous, Gaia Nebbia, Rahul Batra, Jonathan Edgeworth, Samuel C Robson, Angela H Beckett, David M Aanensen, Anthony P Underwood, Corin A Yeats, Khalil Abudahab, Ben E W Taylor, Mirko Menegazzo, Gemma Clark, Wendy Smith, Manjinder Khakh, Vicki M Fleming, Michelle M Lister, Hannah C Howson-Wells, Louise Berry, Tim Boswell, Amelia Joseph, Iona Willingham, Carl Jones, Christopher Holmes, Paul Bird, Thomas Helmer, Karlie Fallon, Julian Tang, Veena Raviprakash, Sharon Campbell, Nicola Sheriff, Victoria Blakey, Lesley-Anne Williams, Matthew W Loose, Nadine Holmes, Christopher Moore, Matthew Carlile, Victoria Wright, Fei Sang, Johnny Debebe, Francesc Coll, Adrian W Signell, Gilberto Betancor, Harry D Wilson, Sahar Eldirdiri, Anita Kenyon, Thomas Davis, Oliver G Pybus, Louis du Plessis, Alex E Zarebski, Jayna Raghwani, Moritz U G Kraemer, Sarah Francois, Stephen W Attwood, Tetyana I Vasylyeva, Marina Escalera Zamudio, Bernardo Gutierrez, M Estee Torok, William L Hamilton, Ian G Goodfellow, Grant Hall, Aminu S Jahun, Yasmin Chaudhry, Myra Hosmillo, Malte L Pinckert, Iliana Georgana, Samuel Moses, Hannah Lowe, Luke Bedford, Jonathan Moore, Susanne Stonehouse, Chloe L Fisher, Ali R Awan, John BoYes, Judith Breuer, Kathryn Ann Harris, Julianne Rose Brown, Divya Shah, Laura Atkinson, Jack C D Lee, Nathaniel Storey, Flavia Flaviani, Adela Alcolea-Medina, Rebecca Williams, Gabrielle Vernet, Michael R Chapman, Lisa J Levett, Judith Heaney, Wendy Chatterton, Monika Pusok, Li Xu-McCrae, Darren L Smith, Matthew Bashton, Gregory R Young, Alison Holmes, Paul Anthony Randell, Alison Cox, Pinglawathee Madona, Frances Bolt, James Price, Siddharth Mookerjee, Manon Ragonnet-Cronin, Fabricia F Nascimento, David Jorgensen, Igor Siveroni, Rob Johnson, Olivia Boyd, Lily Geidelberg, Erik M Volz, Aileen Rowan, Graham P Taylor, Katherine L Smollett, Nicholas J Loman, Joshua Quick, Claire McMurray, Joanne Stockton, Sam Nicholls, Will Rowe, Radoslaw Poplawski, Alan McNally, Rocio T Martinez Nunez, Jenifer Mason, Trevor I Robinson, Elaine O'Toole, Joanne Watts, Cassie Breen, Angela Cowell, Graciela Sluga, Nicholas W Machin, Shazaad S Y Ahmad, Ryan P George, Fenella Halstead, Venkat Sivaprakasam, Wendy Hogsden, Chris J Illingworth, Chris Jackson, Emma C Thomson, James G Shepherd, Patawee Asamaphan, Marc O Niebel, Kathy K Li, Rajiv N Shah, Natasha G Jesudason, Lily Tong, Alice Broos, Daniel Mair, Jenna Nichols, Stephen N Carmichael, Kyriaki Nomikou, Elihu Aranday-Cortes, Natasha Johnson, Igor Starinskij, Ana da Silva Filipe, David L Robertson, Richard J Orton, Joseph Hughes, Sreenu Vattipally, Joshua B Singer, Seema Nickbakhsh, Antony D Hale, Louissa R Macfarlane-Smith, Katherine L Harper, Holli Carden, Yusri Taha, Brendan A I Payne, Shirelle Burton-Fanning, Sheila Waugh, Jennifer Collins, Gary Eltringham, Steven Rushton, Sarah O'Brien, Amanda Bradley, Alasdair Maclean, Guy Mollett, Rachel Blacow, Kate E Templeton, Martin P McHugh, Rebecca Dewar, Elizabeth Wastenge, Samir Dervisevic, Rachael Stanley, Emma J Meader, Lindsay Coupland, Louise Smith, Clive Graham, Edward Barton, Debra Padgett, Garren Scott, Emma Swindells, Jane Greenaway, Andrew Nelson, Clare M McCann, Wen C Yew, Monique Andersson, Timothy Peto, Anita Justice, David Eyre, Derrick Crook, Tim J Sloan, Nichola Duckworth, Sarah Walsh, Anoop J Chauhan, Sharon Glaysher, Kelly Bicknell, Sarah Wyllie, Scott Elliott, Allyson Lloyd, Robert Impey, Nick Levene, Lynn Monaghan, Declan T Bradley, Tim Wyatt, Elias Allara, Clare Pearson, Husam Osman, Andrew Bosworth, Esther Robinson, Peter Muir, Ian B Vipond, Richard Hopes, Hannah M Pymont, Stephanie Hutchings, Martin D Curran, Surendra Parmar, Angie Lackenby, Tamyo Mbisa, Steven Platt, Shahjahan Miah, David Bibby, Carmen Manso, and The COVID-19 Genomics U K (COG-UK) Consortium. Assessing transmissibility of SARS-CoV-2 lineage B.1.1.7 in England. *Nature*, 593(7858):266–269, 2021.

[10] Stefan Elbe and Gemma Buckland-Merrett. Data, disease and diplomacy: Gisaid's innovative contribution to global health. *Global Challenges*, 1(1):33–46, 2017.

[11] Nicholas G Davies, Christopher I Jarvis, Kevin van Zandvoort, Samuel Clifford, Fiona Yueqian Sun, Sebastian Funk, Graham Medley, Yalda Jafari, Sophie R Meakin, Rachel Lowe, Matthew Quaife, Naomi R Waterlow, Rosalind M Eggo, Jiayao Lei, Mihaly Koltai, Fabienne Krauer, Damien C

Tully, James D Munday, Alicia Showering, Anna M Foss, Kiesha Prem, Stefan Flasche, Adam J Kucharski, Sam Abbott, Billy J Quilty, Thibaut Jombart, Alicia Rosello, Gwenan M Knight, Mark Jit, Yang Liu, Jack Williams, Joel Hellewell, Kathleen O'Reilly, Yung-Wai Desmond Chan, Timothy W Russell, Simon R Procter, Akira Endo, Emily S Nightingale, Nikos I Bosse, C Julian Villabona-Arenas, Frank G Sandmann, Amy Gimma, Kaja Abbas, William Waites, Katherine E Atkins, Rosanna C Barnard, Petra Klepac, Hamish P Gibbs, Carl A B Pearson, Oliver Brady, W John Edmunds, Nicholas P Jewell, Karla Diaz-Ordaz, Ruth H Keogh, and CMMID COVID-19 Working Group. Increased mortality in community-tested cases of SARS-CoV-2 lineage B.1.1.7. *Nature*, 593(7858):270–274, 2021.

[12] Robert Challen, Ellen Brooks-Pollock, Jonathan M Read, Louise Dyson, Krasimira Tsaneva-Atanasova, and Leon Danon. Risk of mortality in patients infected with sars-cov-2 variant of concern 202012/1: matched cohort study. *BMJ*, 372, 2021.

[13] Henrik Salje, Cécile Tran Kiem, Noémie Lefrancq, Noémie Courtejoie, Paolo Bosetti, Juliette Paireau, Alessio Andronico, Nathanaël Hozé, Jehanne Richet, Claire-Lise Dubost, Yann Le Strat, Justin Lessler, Daniel Levy-Bruhl, Arnaud Fontanet, Lulla Opatowski, Pierre-Yves Boelle, and Simon Cauchemez. Estimating the burden of SARS-CoV-2 in France. *Science*, 369(6500):208–211, 2020.

[14] Aggiornamento indicazioni sulla Vaccinazione dei soggetti che hanno avuto un'infezione da SARS-CoV-2. `https://redas.services.siag.it/covidArticlesAttachment?attachId=1116555`, 2021.

[15] Hiroshi Nishiura, Tetsuro Kobayashi, Takeshi Miyama, Ayako Suzuki, Sung-mok Jung, Katsuma Hayashi, Ryo Kinoshita, Yichi Yang, Baoyin Yuan, Andrei R Akhmetzhanov, and Natalie M Linton. Estimation of the asymptomatic ratio of novel coronavirus infections (COVID-19). *International Journal of Infectious Diseases*, 94:154–155, may 2020.

[16] Enrico Lavezzo, Elisa Franchin, Constanze Ciavarella, Gina Cuomo-Dannenburg, Luisa Barzon, Claudia Del Vecchio, Lucia Rossi, Riccardo Manganelli, Arianna Loregian, Nicolò Navarin, Davide Abate, Manuela Sciro, Stefano Merigliano, Ettore De Canale, Maria Cristina Vanuzzo, Valeria Besutti, Francesca Saluzzo, Francesco Onelia, Monia Pacenti, Saverio G Parisi, Giovanni Carretta, Daniele Donato, Luciano Flor, Silvia Cocchio, Giulia Masi, Alessandro Sperduti, Lorenzo Cattarino, Renato Salvador, Michele Nicoletti, Federico Caldart, Gioele Castelli, Eleonora Nieddu, Beatrice Labella, Ludovico Fava, Matteo Drigo, Katy A M Gaythorpe, Alessandra R Brazzale, Stefano Toppo, Marta Trevisan, Vincenzo Baldo, Christl A Donnelly, Neil M Ferguson, Ilaria Dorigatti, Andrea Crisanti, Kylie E C Ainslie, Marc Baguelin, Samir Bhatt, Adhiratha Boonyasiri, Olivia Boyd, Lorenzo Cattarino, Constanze Ciavarella, Helen L Coupland, Zulma Cucunubá, Gina Cuomo-Dannenburg, Bimandra A Djafaara, Christl A Donnelly, Ilaria Dorigatti, Sabine L van Elsland, Rich FitzJohn, Seth Flaxman, Katy A M Gaythorpe, Will D Green, Timothy Hallett, Arran Hamlet, David Haw, Natsuko Imai, Benjamin Jeffrey, Edward Knock, Daniel J Laydon, Thomas Mellan, Swapnil Mishra, Gemma Nedjati-Gilani, Pierre Nouvellet, Lucy C Okell, Kris V Parag, Steven Riley, Hayley A Thompson, H Juliette T Unwin, Robert Verity, Michaela A C Vollmer, Patrick G T Walker, Caroline E Walters, Haowei Wang, Yuanrong Wang, Oliver J Watson, Charles Whittaker, Lilith K Whittles, Xiaoyue Xi, Neil M Ferguson, and Imperial College COVID-19 Response Team. Suppression of a SARS-CoV-2 outbreak in the Italian municipality of Vo'. *Nature*, 584(7821):425–429, 2020.

[17] Nicola Perra. Non-pharmaceutical interventions during the covid-19 pandemic: A review. *Physics Reports*, 2021.

[18] Frederik Verelst, Lander Willem, and Philippe Beutels. Behavioural change models for infectious disease transmission: A systematic review (2010-2015). *Journal of The Royal Society Interface*, 13, 12 2016.
